# Comparative effects of sodium glucose cotransporter 2 (SGLT2) inhibitors and dipeptidyl peptidase-4 (DPP4) inhibitors on new-onset atrial fibrillation and stroke outcomes

**DOI:** 10.1101/2021.01.04.21249211

**Authors:** Sharen Lee, Jiandong Zhou, Carlin Chang, Tong Liu, Dong Chang, Wing Tak Wong, Keith SK Leung, Abraham KC Wai, Bernard Man Yung Cheung, Gary Tse, Qingpeng Zhang

## Abstract

**Background:** SGLT2I and DPP4I are medications prescribed for type 2 diabetes mellitus patients. However, there are few population-based studies comparing their effects on incident atrial fibrillation or ischemic stroke.

**Methods:** This was a territory-wide cohort study of type 2 diabetes mellitus patients prescribed SGLT2I or DPP4I between January 1st, 2015 to December 31st, 2019 in Hong Kong. Patients with both DPP4I and SGLT2I use and patients with drug discontinuation were excluded. Patients with prior AF or stroke were excluded for the respective analysis. 1:2 propensity-score matching was conducted for demographics, past comorbidities and medications using nearest-neighbor matching method. Cox models were used to identify significant predictors for new onset heart failure (HF) or myocardial infarction (MI), cardiovascular and all-cause mortality.

**Results:** The AF-free cohort included 49108 patients (mean age: 66.48 years old [SD: 12.89], 55.32% males) and the stroke-free cohort included 49563 patients (27244 males [54.96%], mean baseline age: 66.7 years old [SD: 12.97, max: 104.6 years old]). After propensity score matching, SGLT2i use was associated with a lower risk of new onset AF (HR: 0.43[0.28, 0.66]), cardiovascular mortality (HR: 0.79[0.58, 1.09]) and all-cause mortality (HR: 0.69[0.60, 0.79]) in the AF-free cohort. It was also associated with a lower risk of new onset stroke (0.46[0.33, 0.64]), cardiovascular mortality (HR: 0.74[0.55, 1.00]) and all-cause mortality (HR: 0.64[0.56, 0.74]) in the stroke-free cohort.

**Conclusions:** The novelty of our work si that SGLT2 inhibitors are protective against atrial fibrillation and stroke development for the first time. These findings should be validated in other cohorts.

## Introduction

Type 2 diabetes mellitus is an increasingly prevalent metabolic disease with significant cerebrovascular and cardiovascular complications, including stroke, heart failure (HF) and myocardial infarction. (1) Diabetic patients are associated with a 2.5 times increased risk of ischemic stroke, and the risk is glycemic level-dependent. (2, 3) Atrial fibrillation (AF) is a known risk factor for ischemic stroke, and diabetes mellitus increases the risk of AF through a combination of structural and electrical cardiac remodeling. (4-6) Given the susceptibility of diabetic patients towards cerebrovascular complications, increasing research on the potential cerebrovascular-protective effects of novel anti-diabetic agents has been performed.

Recent studies have shown the superiority of sodium glucose cotransporter-2 inhibitor (SGLT2I) in its ability to lower all-cause and cardiovascular mortality amongst diabetic patients. (7-9) Similarly, dipeptityl peptidase-4 inhibitor (DPP4I) users have been reported to lower the risk of major cardiovascular adverse events, including stroke, though its effect on heart failure remains controversial. (10-12) However, few studies have compared SGLT2I and DPP4I in their effects on stroke and atrial fibrillation. A recent multinational study comparing the cardiovascular outcomes between new SGLT2I and DPP4I users reported lower stroke risk amongst SGLT2I users. (8) However, Hong Kong was not included in its Asia-Pacific data analysis, and the risks of atrial fibrillation have not been examined.

Therefore, to elucidate the cerebrovascular effects of SGLT2I and DPP4I, the present study aims to evaluate the risk of ischemic stroke and atrial fibrillation between SGLT2I and DPP4I users in the Hong Kong population.

## Methods

### Study design and population

This was a retrospective, territory-wide cohort study of type-2 diabetes mellitus patients (n=69521) with SGLT2I/DPP4I use between January 1^st^, 2015 and December 31^st^, 2019 in Hong Kong (**Figure 1**). Patients with both DPP4I and SGLT2I use (n=13215) and patients with drug discontinuation (N=2061) were excluded. Patients with prior AF (n=5137) were excluded from the cohort and a total of 49108 persons fulfilled the eligibility criteria and were included in the AF study cohort for final analysis. Patients with prior stroke (n=4682) were excluded from the cohort and a total of 49563 persons fulfilled the eligibility criteria and were included in the AF study cohort for final analysis. The patients were identified from the Clinical Data Analysis and Reporting System (CDARS), a territory-wide database that centralizes patient information from individual local hospitals to establish comprehensive medical data, including clinical characteristics, disease diagnosis, laboratory results, and drug treatment details. The system has been previously used by both our team and other teams in Hong Kong. (13, 14)

**Figure 1.**
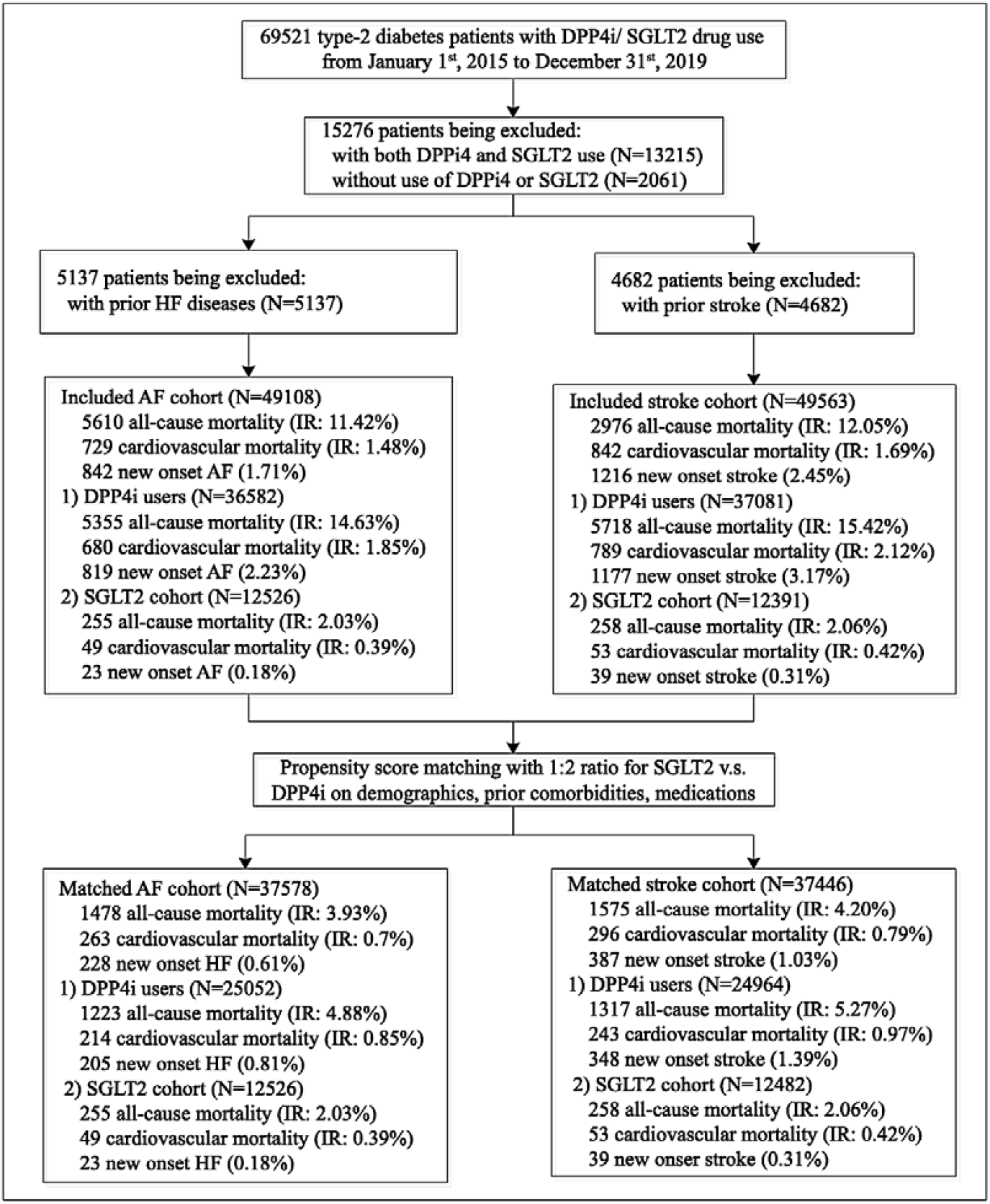
Procedures of data processing for the study cohort.

**Figure 2.**
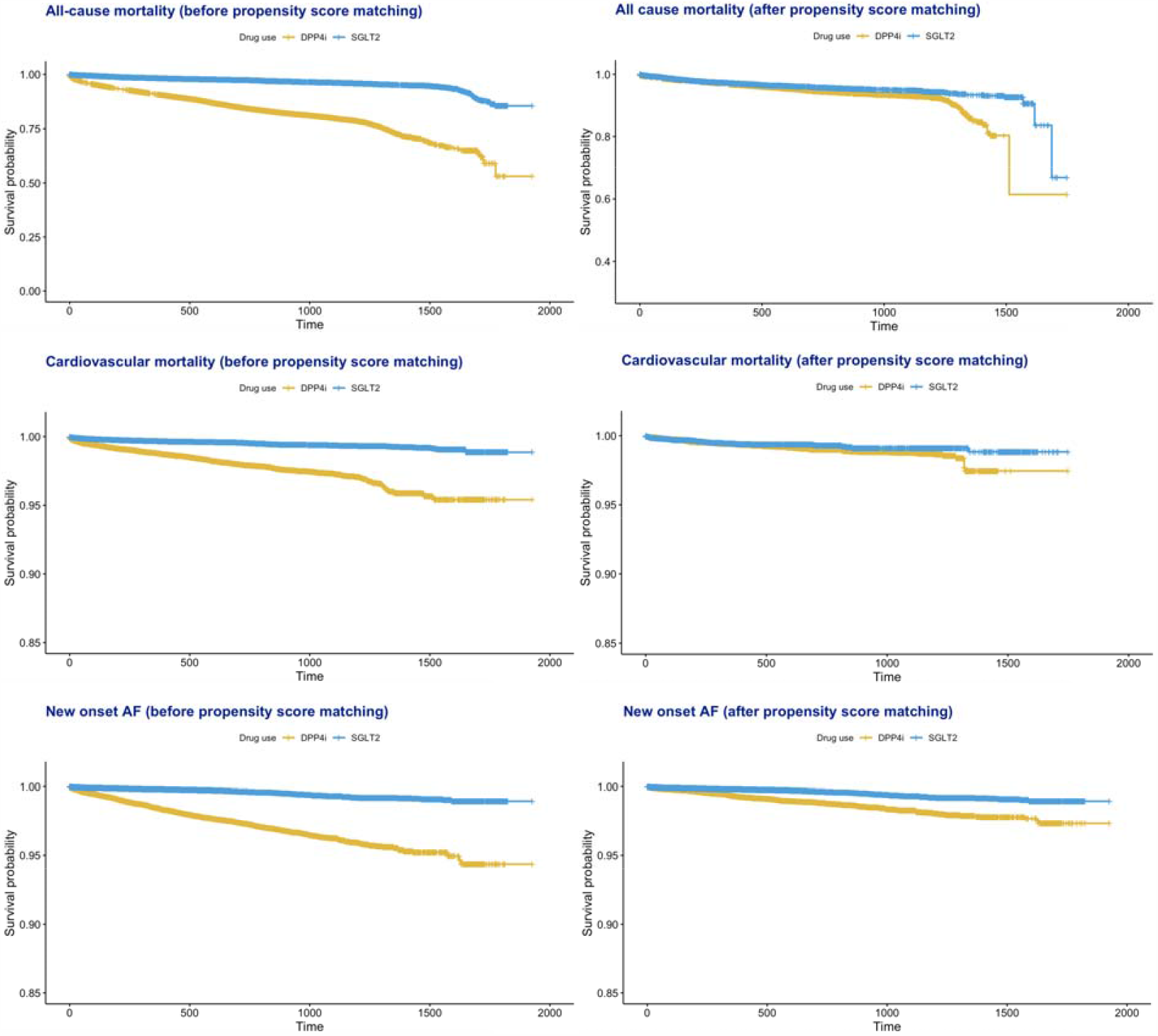
Kaplan-Meier survival curves for all-cause mortality, cardiovascular mortality and new onset AF stratified by drug use of DPP4I and SGLT2I before and after propensity score matching (1:2).

**Figure 3.**
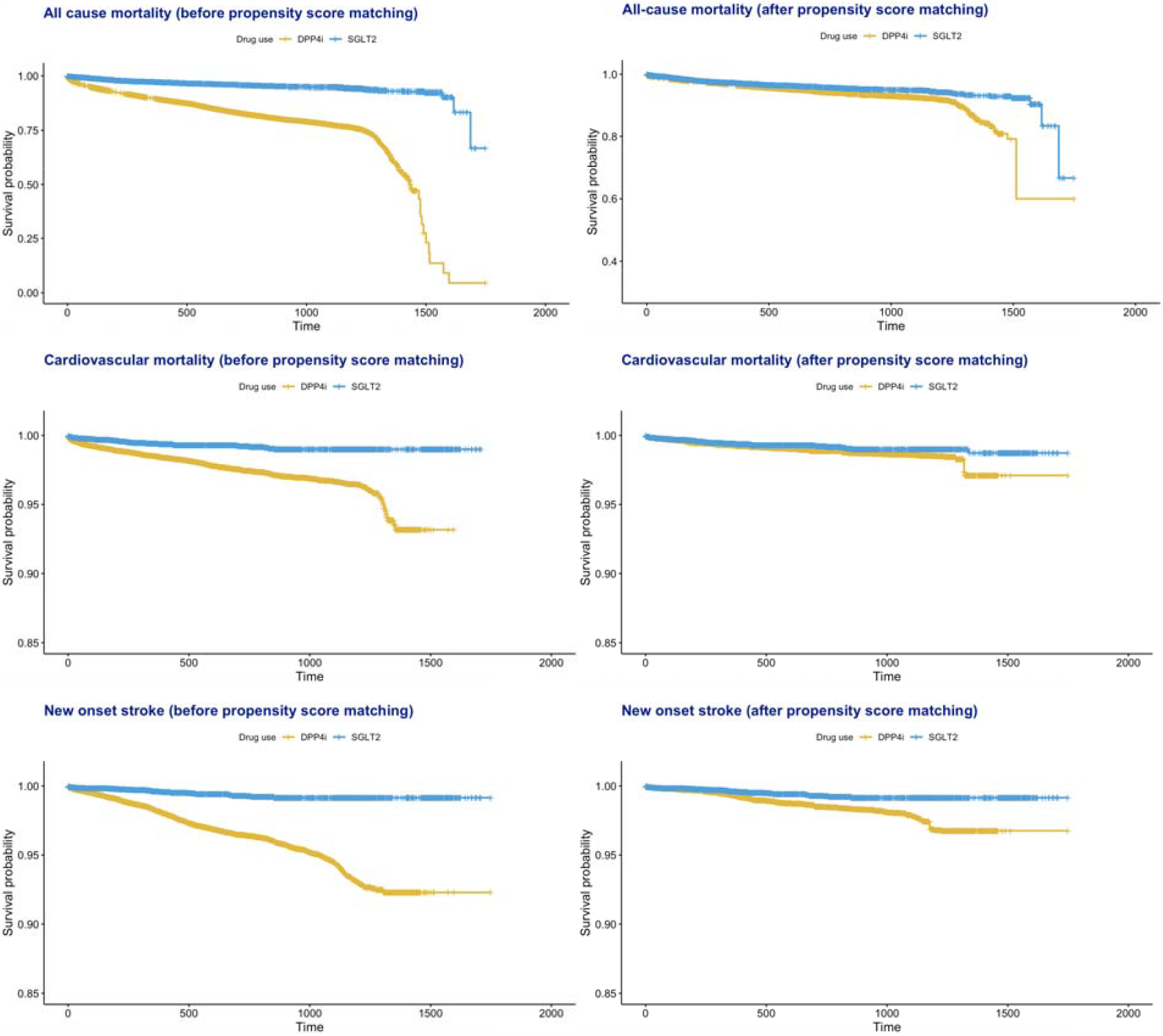
Kaplan-Meier survival curves for all-cause mortality, cardiovascular mortality and new onset stroke stratified by the drug use of DPP4I and SGLT2I before and after propensity score matching (1:2).

Clinical and biochemical data were extracted for the present study. Patients’ demographics include sex and age of initial drug use (baseline). Prior comorbidities before initial drug use were extracted, including cardiovascular, respiratory, renal diseases, endocrine diseases, hypertension, gastrointestinal diseases, HF, ischemic heart disease (IHD), dementia, chronic obstructive pulmonary disease (COPD), stroke and cancer. Charlson comorbidity index was also calculated. Mortality was recorded using the International Classification of Diseases Tenth Edition (ICD-10) coding, whilst the secondary outcomes and comorbidities were documented in CDARS under ICD-9 codes. **Supplementary Table 1** displays the ICD codes used to search for patient outcomes and comorbidities.

Medications histories were also extracted, including gliclazide, glimepiride, metformin, sulphonylurea, insulin, thiazolidinedione, meglitinide, glucagon-like peptide-1 agonist, acarbose, diuretics for hypertension, diuretics for HF, anticoagulants, antiplatelets and lipid-lowering drugs. Baseline laboratory data, defined as complete blood count tests, biochemical tests, and diabetes mellitus tests were extracted. Complete blood count tests include mean corpuscular volume (MCV), basophil, eosinophil, lymphocyte, blast, monocyte, neutrophil, white blood cells, mean corpuscular hemoglobin (MCH), myelocyte, platelet, reticulocyte, red blood cells, and hematocrit (HCT). Biochemical tests include potassium, urate, albumin, sodium, urea, protein, creatinine, alkaline phosphatase (ALP), aspartate transaminase, serum alanine aminotransferase (ALT), and bilirubin. Glycemic and lipid profiles include triglyceride, total cholesterol, low-density lipoprotein (LDL), high-density lipoprotein (HDL), fasting blood glucose, and HbA1c.

### Outcomes and statistical analysis

The primary outcome was new onset ischemic stroke/ transient ischemic attack and AF, and secondary outcomes are all-cause and cardiovascular mortality. Mortality data were obtained from the Hong Kong Death Registry, a population-based official government registry with the registered death records of all Hong Kong citizens linked to CDARS. ICD-10 codes I00-I09, I11, I13, I20-I51 were used to identify cardiovascular mortality. Descriptive statistics are used to summarize baseline clinical and biochemical characteristics of patients with SGLT2I and DPP4I use. For baseline clinical characteristics, the continuous variables were presented as median (95% confidence interval [CI]/ interquartile range [IQR]) or mean (standard deviation [SD]) and the categorical variables were presented as total number (percentage). Continuous variables were compared using the two-tailed Mann-Whitney U test, whilst the two-tailed chi-square test with Yates’ correction was used to test 2×2 contingency data. Univariate Cox regression was used to identify significant predictors for primary and secondary outcomes. Propensity-score matching was performed to generate control for SGLT2I users to compare against DPP4I users in a 1:2 ratio based on age, sex, prior comorbidities, Charlson comorbidity index, medications using nearest-neighbor matching. A standardized mean difference (SMD) of no less than 0.2 between the treatment groups post-weighting was considered negligible. The hazard ratio (HR), 95% CI and P-value were reported. Statistical significance is defined as P-value < 0.05. The statistical analysis was performed with RStudio software (Version: 1.1.456) and Python (Version: 3.6).

## Results

### Basic characteristics of the atrial fibrillation cohort

The AF cohort included 49108 patients (mean age: 66.48 years old [SD: 12.89], 55.32% males). In total, 5610 patients (11.42%) met all-cause mortality outcome after a mean follow up of 519.4 days (SD: 419.82, max: 1748.0), 729(1.48%) passed away due to cardiovascular diseases after a mean follow up of 519.39 days (SD: 419.82, max: 1748.0), and 842 patients (1.71%) developed new AF after a mean follow up of 514.02 days (SD: 418.46, max: 1748.0).

As shown in **Supplementary Table 2**, patients with new onset AF presentation had older baseline age (mean [±SD]: 77.5±10.0; max: 99.7 years old v.s. mean [±SD]:66.3±12.8; max: 104.5 years old, SMD=0.97), and fewer were under 50 years old (0.83% v.s. 9.56%, SMD=0.4), within [50-60] years old (5.46% v.s. 21.73%, SMD=0.49) and [60-70] years old (15.32% v.s. 30.98%, SMD=0.38). More of those with new onset AF were within [70-80] years old (32.06% v.s. 21.53%, SMD=0.24) and older than 80 (46.31% v.s. 16.17%, SMD=0.69). Past comorbidities of cardiovascular diseases (100.00% v.s. 22.50%, SMD=2.62), respiratory diseses (59.26% v.s. 24.12%, SMD=0.76), renal diseases (50.83% v.s. 22.69%, SMD=0.61), hypertension (76.36% v.s. 52.78%, SMD=0.51), gastrointestinal diseases (47.26% v.s. 26.37%, SMD=0.44), and stroke (43.70% v.s. 17.06%, SMD=0.61) were found to be more prevalent in those with new onset AF, and they had a larger Charlson score (mean [±SD]: 3.9±1.7; max: 14.0 v.s. mean [±SD]:2.5±1.7; max: 16.0, SMD=0.82). Patients with new onset AF were more frequently prescribed with DPP4I (97.26% v.s. 74.09%, SMD=0.7; SGLT2I as reference) and antiplatelets (6.55% v.s. 1.06%, SMD=0.29), but were less frequently prescribed lipid-lowering drugs (1.54% v.s. 14.91%, SMD=0.5).

Patients with new onset AF were found to have lower levels of lymphocyte (mean [±SD]: 1.8±0.7; max: 7.5 x10^9/L v.s. mean [±SD]:2.0±1.0; max: 45.4 x10^9/L, SMD=0.25), platelet (mean [±SD]: 217.9±67.7; max: 555.0 x10^9/L v.s. mean [±SD]:239.8±72.9; max: 999.0 x10^9/L, SMD=0.31), red blood cells (mean [±SD]: 4.3±0.7; max: 7.1 x10^12/L v.s. mean [±SD]:4.5±0.7; max: 9.2 x10^12/L, SMD=0.24), urate (mean [±SD]: 0.37±0.1; max: 1.0 mmol/L v.s. mean [±SD]:0.42±0.1; max: 1.3 mmol/L, SMD=0.33), albumin (mean [±SD]: 39.9±4.1; max: 51.0 g/L v.s. mean [±SD]:41.5±4.1; max: 55.1 g/L, SMD=0.39), ALT (mean [±SD]: 22.4±21.3; max: 350.0 U/L v.s. mean [±SD]:28.1±25.1; max: 738.0 U/L, SMD=0.24), and LDL (mean [±SD]: 2.2±0.8; max: 7.1 mmol/L v.s. mean [±SD]:2.4±0.8; max: 17.8 mmol/L, SMD=0.24). However, they had higher levels of HCT (mean [±SD]: 0.41±0.1; max: 0.5 L/L v.s. mean [±SD]:0.38±0.1; max: 0.6 L/L, SMD=0.3), urea (mean [±SD]: 8.5±4.6; max: 48.5 mmol/L v.s. mean [±SD]:6.8±3.8; max: 61.1 mmol/L, SMD=0.4), and creatinine (mean [±SD]: 126.7±103.8; max: 1276.0 umol/L v.s. mean [±SD]:100.3±88.1; max: 1776.0 umol/L, SMD=0.27).

A propensity score matching approach was used to generate an AF control cohort of SGLT2I drug users and DPP4I users with a 1:2 ratio based on demographics, past comorbidities and medications (**Table 1**). After matching, there were more new onset AF patients who were prescribed with diuretics for hypertension (5.13% v.s. 0.00%, SMD=0.33), and diuretics for HF(5.38% v.s. 0.02%, SMD=0.33), but less were prescribed with lipid-lowering drugs (57.56% v.s. 0.23%, SMD=1.63). Those with new onset AF diseases had lower levels of myelocyte (mean [±SD]: 0.29±0.3; max: 1.33 x10^9/L v.s. mean [±SD]:1.08±5.12; max: 36.1 x10^9/L, SMD=0.22), reticulocyte (mean [±SD]: 59.24±23.6; max: 111.0 x10^9/L v.s. mean [±SD]:69.42±29.78; max: 175.0 x10^9/L, SMD=0.38), urate (mean [±SD]: 0.37±0.1; max: 0.91 mmol/L v.s. mean [±SD]:0.41±0.11; max: 0.97 mmol/L, SMD=0.31), creatinine (mean [±SD]: 77.36±21.55; max: 757.0 umol/L v.s. mean [±SD]:91.32±57.29; max: 1294.0 umol/L, SMD=0.32) and urea (mean [±SD]: 5.58±1.77; max: 44.6 mmol/L v.s. mean [±SD]:6.36±2.97; max: 43.21 mmol/L, SMD=0.32). However, they had higher levels of red blood cells (mean [±SD]: 4.77±0.59; max: 9.18 x10^12/L v.s. mean [±SD]:4.61±0.64; max: 7.86 x10^12/L, SMD=0.26), HCT (mean [±SD]: 0.41±0.04; max: 0.61 L/L v.s. mean [±SD]:0.4±0.05; max: 0.57 L/L, SMD=0.31), and ALT (mean [±SD]: 34.09±30.81; max: 738.0 U/L v.s. mean [±SD]:28.14±23.77; max: 605.0 U/L, SMD=0.22).

**Table 1.**
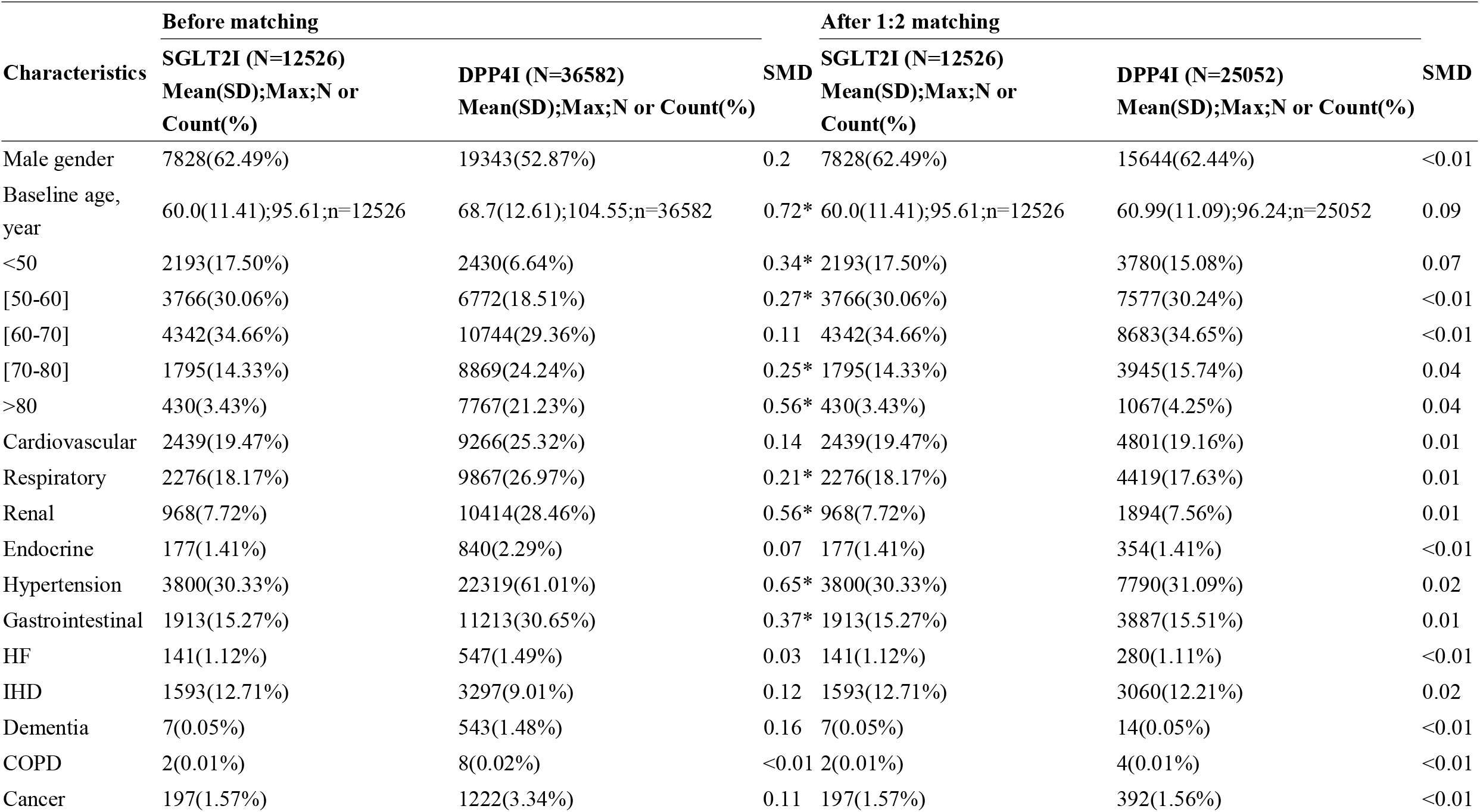
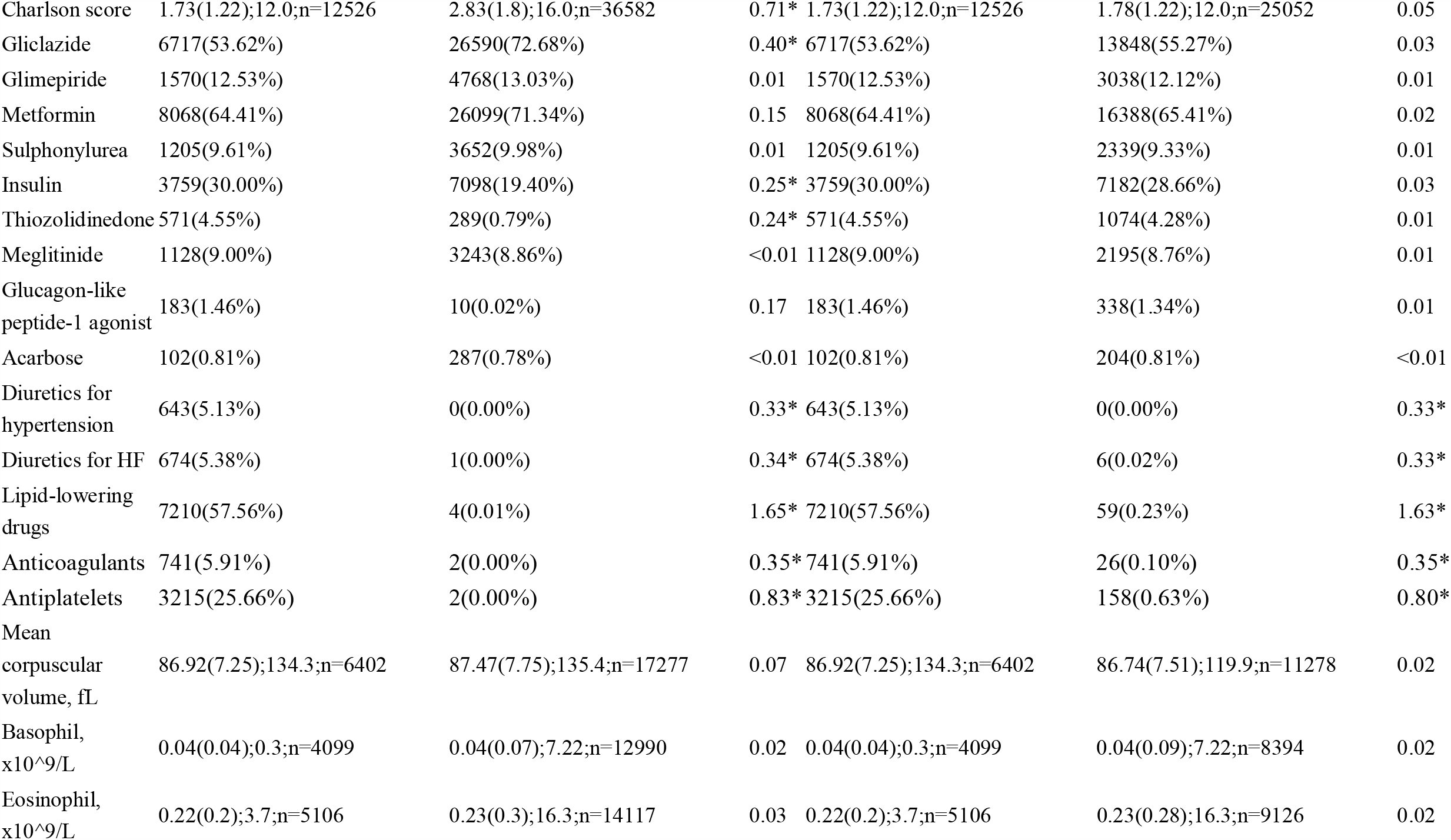
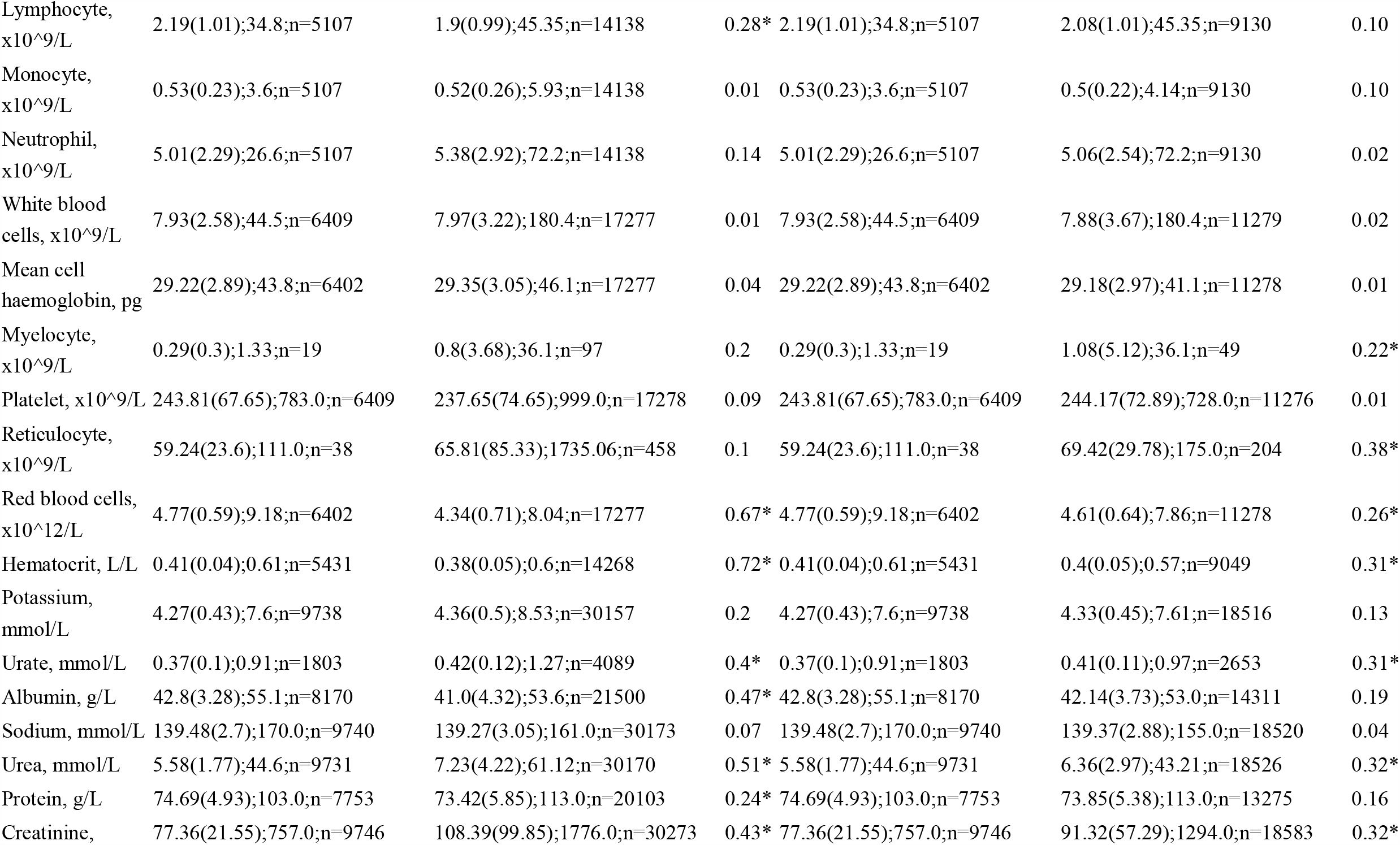
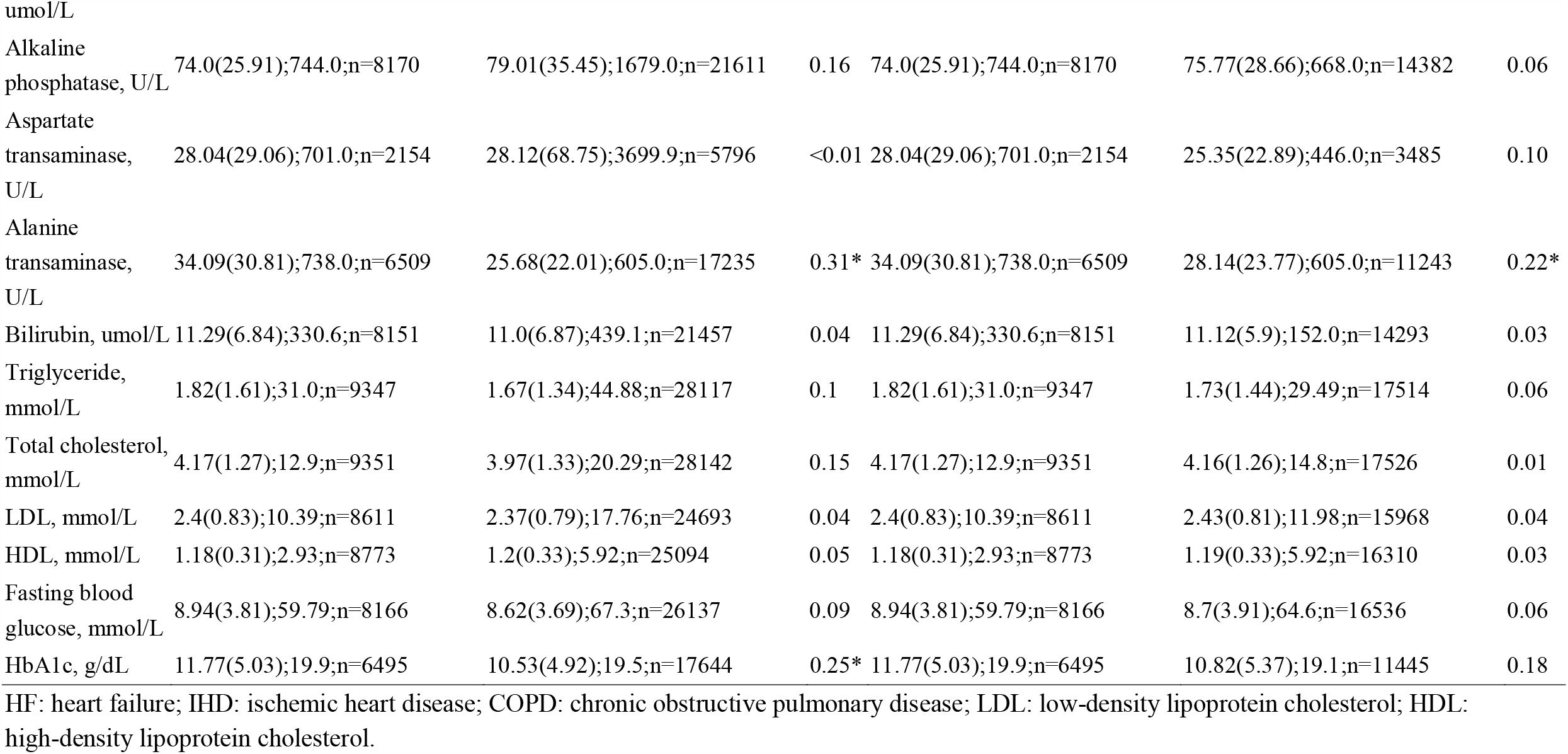
Baseline characteristics of patients with DPP4I or SGLT2I use before and after propensity score matching (1:2) in the AF cohort. * for SMD>0.2

### Basic characteristics of stroke cohort

The stroke cohort included 49563 patients (27244 males [54.96%], mean baseline age: 66.7 years old [SD: 12.97, max: 104.6 years old]). Overall 5976 patients (12.05%) passed after a mean follow up of 518.1 days (SD: 418.94, max: 1748 days), 842 patients (1.69%) passed away from cardiovascular diseases after a mean follow up of 518.1 days (SD: 418.9, max: 1748 days), and 1216 patients (2.45%) developed new onset stroke after mean follow up of 512.1 days (SD: 417.2, max: 1748 days).

As shown in **Supplementary Table 3**, patients with new onset stroke had older baseline age (mean [±SD]: 74.2±10.6; max: 102.3 years old v.s. mean [±SD]:66.6±13.0; max: 104.5 years old, SMD=0.65): less under 50 years old (1.80% v.s. 9.45%, SMD=0.34), within [50-60] years old (8.22% v.s. 21.37%, SMD=0.38) but more within [70-80] years old (33.05% v.s. 21.65%, SMD=0.26) and over 80 years old (33.14% v.s. 17.09%, SMD=0.38). More new onset stroke patients had prior comorbidities of cardiovascular diseases (37.82% v.s. 26.13%, SMD=0.25), respiratory diseases (37.99% v.s. 25.34%, SMD=0.27), renal diseases (37.41% v.s. 23.59%, SMD=0.3), hypertension (85.03% v.s. 52.41%, SMD=0.75), gastrointestinal diseases (38.81% v.s. 27.02%, SMD=0.25) and had a larger Charlson score (mean [±SD]: 3.4±1.6; max: 14.0 v.s. mean [±SD]:2.5±1.8; max: 15.0, SMD=0.48).

DPP4I (96.79% v.s. 74.26%, SMD=0.68), gliclazide (78.78% v.s. 67.47%, SMD=0.26) and antiplatelets (6.25% v.s. 1.48%, SMD=0.25) were more frequently prescribed for those with new onset stroke, whilst less commonly prescribed with lipid-lowering drugs (2.54% v.s. 14.71%, SMD=0.44). In addition, patients with new onset stroke were found to have lower levels of red blood cells (mean [±SD]: 4.3±0.7; max: 6.6 x10^12/L v.s. mean [±SD]:4.4±0.7; max: 9.2 x10^12/L, SMD=0.25) and albumin (mean [±SD]: 40.5±4.2; max: 53.1 g/L v.s. mean [±SD]:41.5±4.2; max: 55.1 g/L, SMD=0.22), but have higher HCT (mean [±SD]: 0.41±0.1; max: 0.5 L/L v.s. mean [±SD]:0.4±0.1; max: 0.6 L/L, SMD=0.28).

A propensity score matching approach was used to generate a stroke control cohort of SGLT2I drug users and DPP4I users with 1:2 ratio based on demographics, past comorbidities and medications (**Table 2**). After matching, there were more new stroke patients who were prescribed with diuretics for hypertension (5.04% v.s. 0.00%, SMD=0.33), diuretics for HF (5.92% v.s. 0.01%, SMD=0.35), anticoagulants (6.63% v.s. 0.08%, SMD=0.37), antiplatelets (24.80% v.s. 0.62%, SMD=0.78) and lipid-lowering drugs (57.19% v.s. 0.22%, SMD=1.62).

**Table 2.**
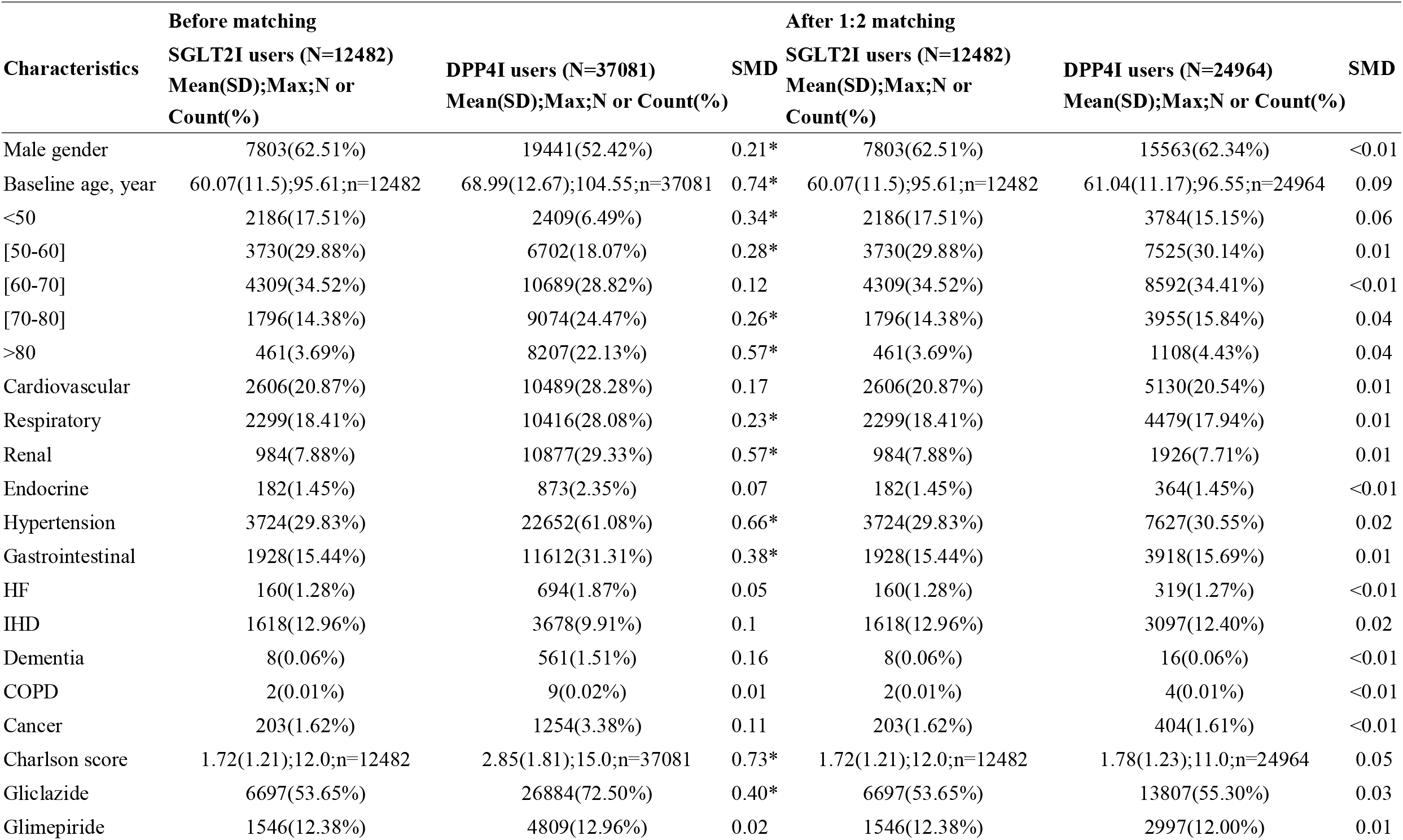
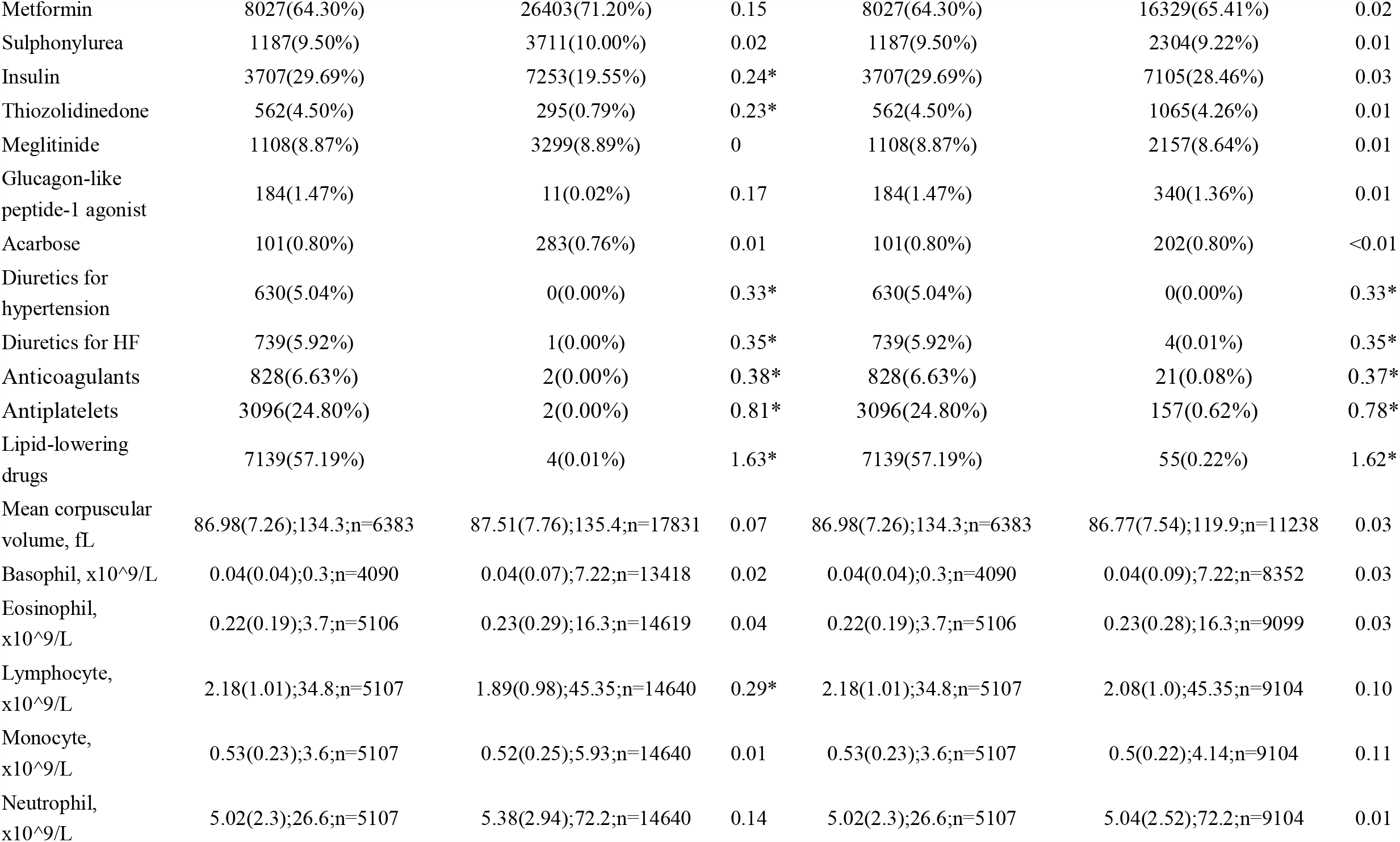
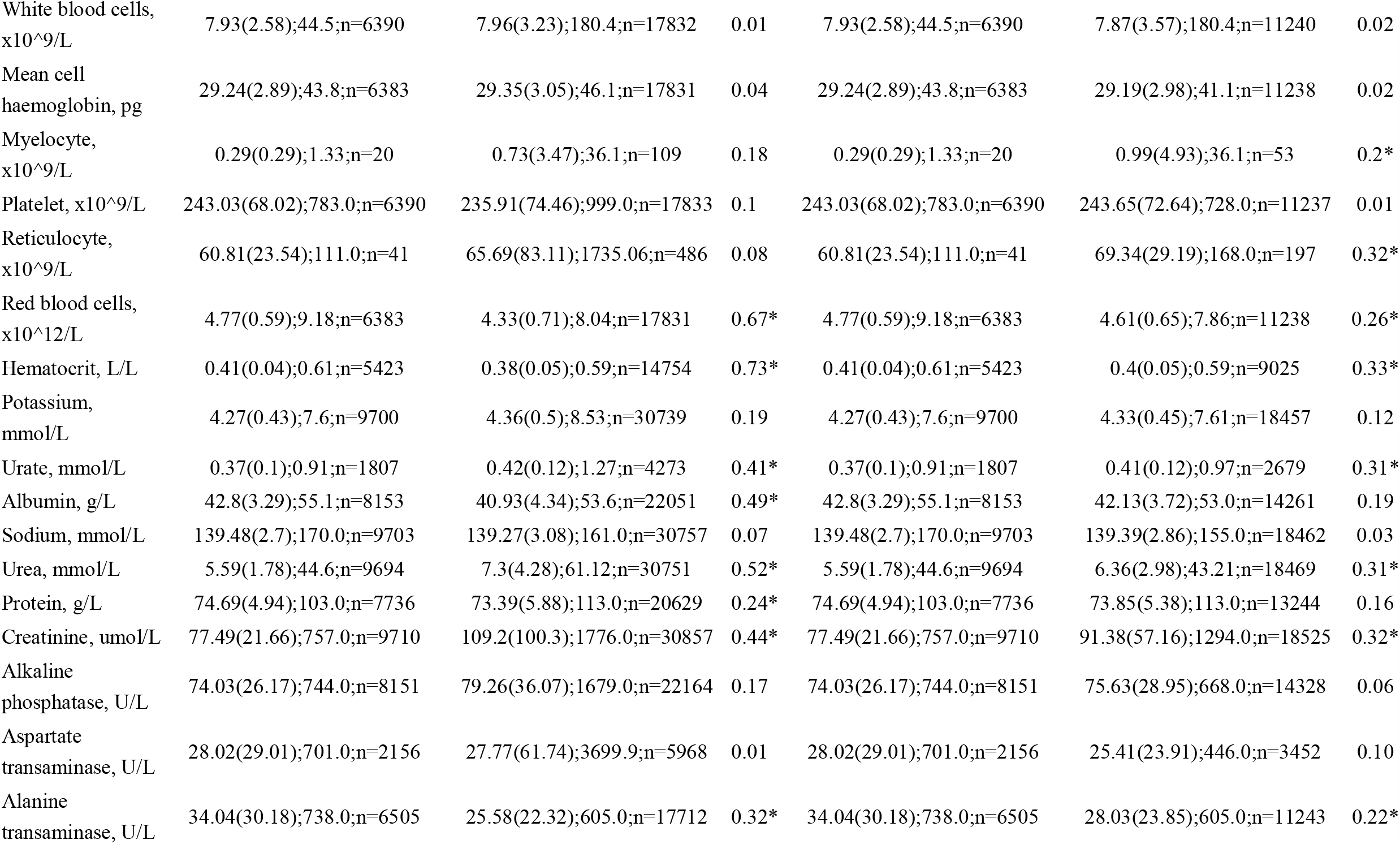
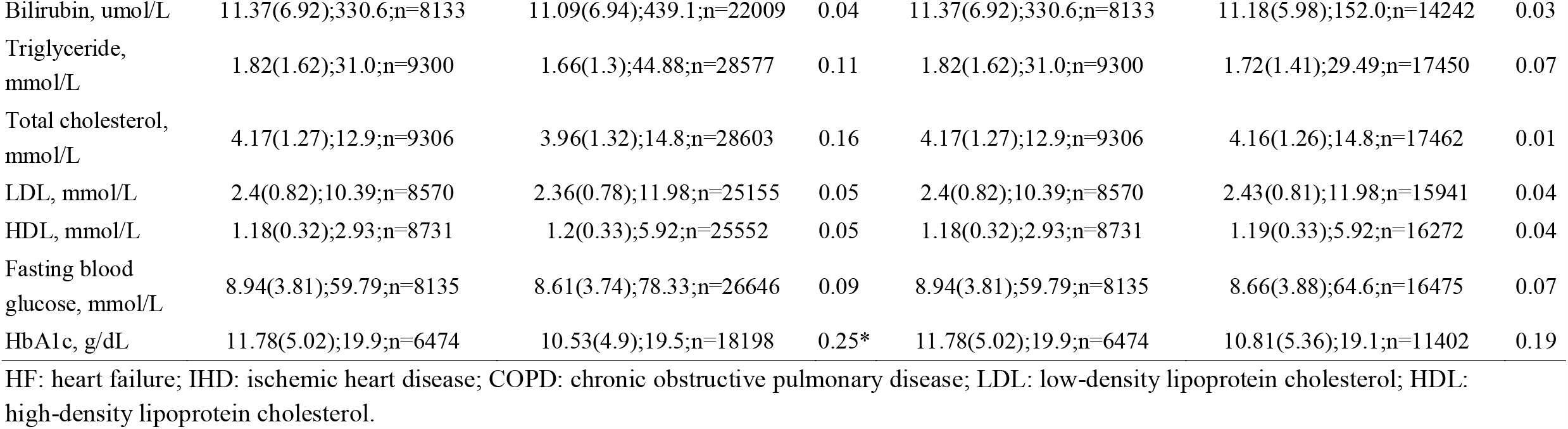
Baseline characteristics of patients with DPP4I or SGLT2I use before and after propensity score matching (1:2) in the stroke cohort. * for SMD>0.2

| Characteristics | Before matching |  |  | After 1:2 matching |  |  |
| --- | --- | --- | --- | --- | --- | --- |
|  | SGLT2I users (N=12482)<br>Mean(SD);Max;N or<br>Count(%) | DPP4I users (N=37081)<br>Mean(SD);Max;N or<br>Count(%) | SMD | SGLT2I users (N=12482)<br>Mean(SD);Max;N or<br>Count(%) | DPP4I users (N=24964)<br>Mean(SD);Max;N or<br>Count(%) | SMD |
| Male gender | 7803(62.51%) | 19441(52.42%) | 0.21* | 7803(62.51%) | 15563(62.34%) | <0.01 |
| Baseline age, year | 60.07(11.5);95.61;n=12482 | 68.99(12.67);104.55;n=37081 | 0.74* | 60.07(11.5);95.61;n=12482 | 61.04(11.17);96.55;n=24964 | 0.09 |
| <50 | 2186(17.51%) | 2409(6.49%) | 0.34* | 2186(17.51%) | 3784(15.15%) | 0.06 |
| [50-60] | 3730(29.88%) | 6702(18.07%) | 0.28* | 3730(29.88%) | 7525(30.14%) | 0.01 |
| [60-70] | 4309(34.52%) | 10689(28.82%) | 0.12 | 4309(34.52%) | 8592(34.41%) | <0.01 |
| [70-80] | 1796(14.38%) | 9074(24.47%) | 0.26* | 1796(14.38%) | 3955(15.84%) | 0.04 |
| >80 | 461(3.69%) | 8207(22.13%) | 0.57* | 461(3.69%) | 1108(4.43%) | 0.04 |
| Cardiovascular | 2606(20.87%) | 10489(28.28%) | 0.17 | 2606(20.87%) | 5130(20.54%) | 0.01 |
| Respiratory | 2299(18.41%) | 10416(28.08%) | 0.23* | 2299(18.41%) | 4479(17.94%) | 0.01 |
| Renal | 984(7.88%) | 10877(29.33%) | 0.57* | 984(7.88%) | 1926(7.71%) | 0.01 |
| Endocrine | 182(1.45%) | 873(2.35%) | 0.07 | 182(1.45%) | 364(1.45%) | <0.01 |
| Hypertension | 3724(29.83%) | 22652(61.08%) | 0.66* | 3724(29.83%) | 7627(30.55%) | 0.02 |
| Gastrointestinal | 1928(15.44%) | 11612(31.31%) | 0.38* | 1928(15.44%) | 3918(15.69%) | 0.01 |
| HF | 160(1.28%) | 694(1.87%) | 0.05 | 160(1.28%) | 319(1.27%) | <0.01 |
| IHD | 1618(12.96%) | 3678(9.91%) | 0.1 | 1618(12.96%) | 3097(12.40%) | 0.02 |
| Dementia | 8(0.06%) | 561(1.51%) | 0.16 | 8(0.06%) | 16(0.06%) | <0.01 |
| COPD | 2(0.01%) | 9(0.02%) | 0.01 | 2(0.01%) | 4(0.01%) | <0.01 |
| Cancer | 203(1.62%) | 1254(3.38%) | 0.11 | 203(1.62%) | 404(1.61%) | <0.01 |
| Charlson score | 1.72(1.21);12.0;n=12482 | 2.85(1.81);15.0;n=37081 | 0.73* | 1.72(1.21);12.0;n=12482 | 1.78(1.23);11.0;n=24964 | 0.05 |
| Gliclazide | 6697(53.65%) | 26884(72.50%) | 0.40* | 6697(53.65%) | 13807(55.30%) | 0.03 |
| Glimepiride | 1546(12.38%) | 4809(12.96%) | 0.02 | 1546(12.38%) | 2997(12.00%) | 0.01 |
| Metformin | 8027(64.30%) | 26403(71.20%) | 0.15 | 8027(64.30%) | 16329(65.41%) | 0.02 |
| Sulphonylurea | 1187(9.50%) | 3711(10.00%) | 0.02 | 1187(9.50%) | 2304(9.22%) | 0.01 |
| Insulin | 3707(29.69%) | 7253(19.55%) | 0.24* | 3707(29.69%) | 7105(28.46%) | 0.03 |
| Thiozolidinedone | 562(4.50%) | 295(0.79%) | 0.23* | 562(4.50%) | 1065(4.26%) | 0.01 |
| Meglitinide | 1108(8.87%) | 3299(8.89%) | 0 | 1108(8.87%) | 2157(8.64%) | 0.01 |
| Glucagon-like<br>peptide-1 agonist | 184(1.47%) | 11(0.02%) | 0.17 | 184(1.47%) | 340(1.36%) | 0.01 |
| Acarbose | 101(0.80%) | 283(0.76%) | 0.01 | 101(0.80%) | 202(0.80%) | <0.01 |
| Diuretics for<br>hypertension | 630(5.04%) | 0(0.00%) | 0.33* | 630(5.04%) | 0(0.00%) | 0.33* |
| Diuretics for HF | 739(5.92%) | 1(0.00%) | 0.35* | 739(5.92%) | 4(0.01%) | 0.35* |
| Anticoagulants | 828(6.63%) | 2(0.00%) | 0.38* | 828(6.63%) | 21(0.08%) | 0.37* |
| Antiplatelets | 3096(24.80%) | 2(0.00%) | 0.81* | 3096(24.80%) | 157(0.62%) | 0.78* |
| Lipid-lowering<br>drugs | 7139(57.19%) | 4(0.01%) | 1.63* | 7139(57.19%) | 55(0.22%) | 1.62* |
| Mean corpuscular<br>volume, fL | 86.98(7.26);134.3;n=6383 | 87.51(7.76);135.4;n=17831 | 0.07 | 86.98(7.26);134.3;n=6383 | 86.77(7.54);119.9;n=11238 | 0.03 |
| Basophil, x10<sup>9</sup>/L | 0.04(0.04);0.3;n=4090 | 0.04(0.07);7.22;n=13418 | 0.02 | 0.04(0.04);0.3;n=4090 | 0.04(0.09);7.22;n=8352 | 0.03 |
| Eosinophil,<br>x10<sup>9</sup>/L | 0.22(0.19);3.7;n=5106 | 0.23(0.29);16.3;n=14619 | 0.04 | 0.22(0.19);3.7;n=5106 | 0.23(0.28);16.3;n=9099 | 0.03 |
| Lymphocyte,<br>x10<sup>9</sup>/L | 2.18(1.01);34.8;n=5107 | 1.89(0.98);45.35;n=14640 | 0.29* | 2.18(1.01);34.8;n=5107 | 2.08(1.0);45.35;n=9104 | 0.10 |
| Monocyte,<br>x10<sup>9</sup>/L | 0.53(0.23);3.6;n=5107 | 0.52(0.25);5.93;n=14640 | 0.01 | 0.53(0.23);3.6;n=5107 | 0.5(0.22);4.14;n=9104 | 0.11 |
| Neutrophil,<br>x10<sup>9</sup>/L | 5.02(2.3);26.6;n=5107 | 5.38(2.94);72.2;n=14640 | 0.14 | 5.02(2.3);26.6;n=5107 | 5.04(2.52);72.2;n=9104 | 0.01 |
| White blood cells,<br>x10<sup>9</sup>/L | 7.93(2.58);44.5;n=6390 | 7.96(3.23);180.4;n=17832 | 0.01 | 7.93(2.58);44.5;n=6390 | 7.87(3.57);180.4;n=11240 | 0.02 |
| Mean cell<br>haemoglobin, pg | 29.24(2.89);43.8;n=6383 | 29.35(3.05);46.1;n=17831 | 0.04 | 29.24(2.89);43.8;n=6383 | 29.19(2.98);41.1;n=11238 | 0.02 |
| Myelocyte,<br>x10<sup>9</sup>/L | 0.29(0.29);1.33;n=20 | 0.73(3.47);36.1;n=109 | 0.18 | 0.29(0.29);1.33;n=20 | 0.99(4.93);36.1;n=53 | 0.2* |
| Platelet, x10<sup>9</sup>/L | 243.03(68.02);783.0;n=6390 | 235.91(74.46);999.0;n=17833 | 0.1 | 243.03(68.02);783.0;n=6390 | 243.65(72.64);728.0;n=11237 | 0.01 |
| Reticulocyte,<br>x10<sup>9</sup>/L | 60.81(23.54);111.0;n=41 | 65.69(83.11);1735.06;n=486 | 0.08 | 60.81(23.54);111.0;n=41 | 69.34(29.19);168.0;n=197 | 0.32* |
| Red blood cells,<br>x10<sup>12</sup>/L | 4.77(0.59);9.18;n=6383 | 4.33(0.71);8.04;n=17831 | 0.67* | 4.77(0.59);9.18;n=6383 | 4.61(0.65);7.86;n=11238 | 0.26* |
| Hematocrit, L/L | 0.41(0.04);0.61;n=5423 | 0.38(0.05);0.59;n=14754 | 0.73* | 0.41(0.04);0.61;n=5423 | 0.4(0.05);0.59;n=9025 | 0.33* |
| Potassium,<br>mmol/L | 4.27(0.43);7.6;n=9700 | 4.36(0.5);8.53;n=30739 | 0.19 | 4.27(0.43);7.6;n=9700 | 4.33(0.45);7.61;n=18457 | 0.12 |
| Urate, mmol/L | 0.37(0.1);0.91;n=1807 | 0.42(0.12);1.27;n=4273 | 0.41* | 0.37(0.1);0.91;n=1807 | 0.41(0.12);0.97;n=2679 | 0.31* |
| Albumin, g/L | 42.8(3.29);55.1;n=8153 | 40.93(4.34);53.6;n=22051 | 0.49* | 42.8(3.29);55.1;n=8153 | 42.13(3.72);53.0;n=14261 | 0.19 |
| Sodium, mmol/L | 139.48(2.7);170.0;n=9703 | 139.27(3.08);161.0;n=30757 | 0.07 | 139.48(2.7);170.0;n=9703 | 139.39(2.86);155.0;n=18462 | 0.03 |
| Urea, mmol/L | 5.59(1.78);44.6;n=9694 | 7.3(4.28);61.12;n=30751 | 0.52* | 5.59(1.78);44.6;n=9694 | 6.36(2.98);43.21;n=18469 | 0.31* |
| Protein, g/L | 74.69(4.94);103.0;n=7736 | 73.39(5.88);113.0;n=20629 | 0.24* | 74.69(4.94);103.0;n=7736 | 73.85(5.38);113.0;n=13244 | 0.16 |
| Creatinine, umol/L | 77.49(21.66);757.0;n=9710 | 109.2(100.3);1776.0;n=30857 | 0.44* | 77.49(21.66);757.0;n=9710 | 91.38(57.16);1294.0;n=18525 | 0.32* |
| Alkaline<br>phosphatase, U/L | 74.03(26.17);744.0;n=8151 | 79.26(36.07);1679.0;n=22164 | 0.17 | 74.03(26.17);744.0;n=8151 | 75.63(28.95);668.0;n=14328 | 0.06 |
| Aspartate<br>transaminase, U/L | 28.02(29.01);701.0;n=2156 | 27.77(61.74);3699.9;n=5968 | 0.01 | 28.02(29.01);701.0;n=2156 | 25.41(23.91);446.0;n=3452 | 0.10 |
| Alanine<br>transaminase, U/L | 34.04(30.18);738.0;n=6505 | 25.58(22.32);605.0;n=17712 | 0.32* | 34.04(30.18);738.0;n=6505 | 28.03(23.85);605.0;n=11243 | 0.22* |
| Bilirubin, umol/L | 11.37(6.92);330.6;n=8133 | 11.09(6.94);439.1;n=22009 | 0.04 | 11.37(6.92);330.6;n=8133 | 11.18(5.98);152.0;n=14242 | 0.03 |
| Triglyceride, mmol/L | 1.82(1.62);31.0;n=9300 | 1.66(1.3);44.88;n=28577 | 0.11 | 1.82(1.62);31.0;n=9300 | 1.72(1.41);29.49;n=17450 | 0.07 |
| Total cholesterol, mmol/L | 4.17(1.27);12.9;n=9306 | 3.96(1.32);14.8;n=28603 | 0.16 | 4.17(1.27);12.9;n=9306 | 4.16(1.26);14.8;n=17462 | 0.01 |
| LDL, mmol/L | 2.4(0.82);10.39;n=8570 | 2.36(0.78);11.98;n=25155 | 0.05 | 2.4(0.82);10.39;n=8570 | 2.43(0.81);11.98;n=15941 | 0.04 |
| HDL, mmol/L | 1.18(0.32);2.93;n=8731 | 1.2(0.33);5.92;n=25552 | 0.05 | 1.18(0.32);2.93;n=8731 | 1.19(0.33);5.92;n=16272 | 0.04 |
| Fasting blood glucose, mmol/L | 8.94(3.81);59.79;n=8135 | 8.61(3.74);78.33;n=26646 | 0.09 | 8.94(3.81);59.79;n=8135 | 8.66(3.88);64.6;n=16475 | 0.07 |
| HbA1c, g/dL | 11.78(5.02);19.9;n=6474 | 10.53(4.9);19.5;n=18198 | 0.25* | 11.78(5.02);19.9;n=6474 | 10.81(5.36);19.1;n=11402 | 0.19 |

HF: heart failure; IHD: ischemic heart disease; COPD: chronic obstructive pulmonary disease; LDL: low-density lipoprotein cholesterol; HDL: high-density lipoprotein cholesterol.

**Table 3.** Cox-proportional hazard model analysis for all-cause mortality, cardiovascular mortality and new onset AF after propensity score matching (1:2). * for p≤ 0.05, ** for p ≤ 0.01, *** for p ≤ 0.001

| Characteristics | All-cause mortality<br>HR [95% CI] | P value | Cardiovascular mortality<br>HR [95% CI] | P value | New onset AF<br>HR [95% CI] | P value |
| --- | --- | --- | --- | --- | --- | --- |
| Male gender | 1.36[1.22, 1.51] | <0.0001*** | 1.18[0.91, 1.52] | 0.2100 | 1.47[1.11, 1.95] | 0.0080** |
| Baseline age, year | 1.08[1.07, 1.08] | <0.0001*** | 1.07[1.06, 1.08] | <0.0001*** | 1.09[1.08, 1.11] | <0.0001*** |
| <50 | 0.28[0.22, 0.35] | <0.0001*** | 0.34[0.21, 0.56] | <0.0001*** | 0.11[0.05, 0.27] | <0.0001*** |
| [50-60] | 0.53[0.47, 0.60] | <0.0001*** | 0.58[0.43, 0.79] | 0.0004*** | 0.43[0.31, 0.61] | <0.0001*** |
| [60-70] | 0.77[0.68, 0.86] | <0.0001*** | 0.70[0.53, 0.92] | 0.0108* | 0.78[0.58, 1.04] | 0.0876. |
| [70-80] | 2.55[2.28, 2.85] | <0.0001*** | 2.59[1.99, 3.37] | <0.0001*** | 3.43[2.61, 4.49] | <0.0001*** |
| >80 | 5.79[5.07, 6.63] | <0.0001*** | 5.07[3.65, 7.06] | <0.0001*** | 5.80[4.11, 8.19] | <0.0001*** |
| Cardiovascular | 4.4[3.9, 4.8] | <0.0001*** | 15.3[11.5, 20.4] | <0.0001*** | - | - |
| Respiratory | 6.21[5.60, 6.88] | <0.0001*** | 5.80[4.55, 7.39] | <0.0001*** | 2.57[1.96, 3.37] | <0.0001*** |
| Renal | 3.40[3.01, 3.85] | <0.0001*** | 3.54[2.65, 4.72] | <0.0001*** | 1.93[1.33, 2.80] | 0.0006*** |
| Endocrine | 3.56[2.83, 4.47] | <0.0001*** | 0.50[0.12, 2.01] | 0.3280 | 2.99[1.59, 5.64] | 0.0007*** |
| Hypertension | 1.90[1.72, 2.11] | <0.0001*** | 1.74[1.36, 2.22] | <0.0001*** | 3.52[2.70, 4.60] | <0.0001*** |
| Gastrointestinal | 2.51[2.25, 2.80] | <0.0001*** | 0.85[0.60, 1.21] | 0.3670 | 1.99[1.49, 2.66] | <0.0001*** |
| HF | 6.62[5.29, 8.28] | <0.0001*** | 8.19[5.07, 13.21] | <0.0001*** | 8.63[5.11, 14.57] | <0.0001*** |
| IHD | 2.55[2.26, 2.88] | <0.0001*** | 6.41[5.02, 8.18] | <0.0001*** | 2.27[1.65, 3.14] | <0.0001*** |
| Dementia | 6.73[2.52, 17.97] | 0.0001*** | - | - | - | - |
| COPD | 5.27[0.74, 37.46] | 0.0966. | - | - | - | - |
| Cancer | 5.37[4.37, 6.61] | <0.0001*** | 1.17[0.44, 3.14] | 0.7560 | 5.30[3.09, 9.10] | <0.0001*** |
| Charlson score | 1.75[1.71, 1.79] | <0.0001*** | 1.68[1.59, 1.79] | <0.0001*** | 1.75[1.64, 1.87] | <0.0001*** |
| SGLT2I | 0.69[0.60, 0.79] | <0.0001*** | 0.79[0.58, 1.09] | 0.1510 | 0.43[0.28, 0.66] | 0.0001*** |
| DPP4I | 1.45[1.27, 1.67] | <0.0001*** | 1.26[0.92, 1.72] | 0.1510 | 2.34[1.51, 3.60] | 0.0001*** |
| Gliclazide | 1.35[1.22, 1.50] | <0.0001*** | 1.93[1.49, 2.50] | <0.0001*** | 2.12[1.60, 2.82] | <0.0001*** |
| Glimepiride | 0.83[0.70, 0.97] | 0.0226* | 0.87[0.59, 1.27] | 0.4570 | 0.81[0.54, 1.23] | 0.3330 |
| Metformin | 1.00[0.89, 1.11] | 0.9310 | 1.00[0.77, 1.29] | 0.9760 | 1.41[1.04, 1.89] | 0.0248* |
| Sulphonylurea | 0.79[0.66, 0.95] | 0.0105* | 0.75[0.48, 1.17] | 0.2000 | 0.71[0.44, 1.15] | 0.1640 |
| Insulin | 1.05[0.94, 1.17] | 0.3710 | 1.13[0.88, 1.46] | 0.3410 | 0.48[0.34, 0.66] | <0.0001*** |
| Thiozolidinedone | 0.52[0.39, 0.69] | <0.0001*** | 0.65[0.34, 1.22] | 0.1820 | 0.34[0.14, 0.84] | 0.0187* |
| Meglitinide | 0.82[0.68, 0.99] | 0.0348* | 0.79[0.51, 1.24] | 0.3080 | 0.71[0.43, 1.16] | 0.1710 |
| Glucagon-like peptide-1 agonist | 0.61[0.39, 0.95] | 0.0281* | 0.19[0.03, 1.32] | 0.0926. | 0.00[0.00, Inf] | 0.9900 |
| Acarbose | 1.66[1.06, 2.57] | 0.0253* | 0.94[0.23, 3.76] | 0.9260 | 4.51[2.23, 9.14] | <0.0001*** |
| Diuretics for hypertension | 0.65[0.38, 1.10] | 0.1060 | 0.28[0.04, 1.96] | 0.1980 | 0.99[0.32, 3.10] | 0.9870 |
| Diuretics for HF | 1.95[1.41, 2.69] | 0.0001*** | 5.41[3.30, 8.86] | <0.0001*** | 0.36[0.05, 2.58] | 0.3110 |
| Anticoagulants | 1.39[0.97, 1.99] | 0.0719. | 4.27[2.53, 7.19] | <0.0001*** | 1.67[0.62, 4.49] | 0.313 |
| Antiplatelets | 0.99[0.81, 1.21] | 0.918 | 1.76[1.2, 2.57] | 0.0039** | 0.72[0.37, 1.42] | 0.0345 |
| Lipid-lowering drugs | 0.65[0.54, 0.77] | <0.0001*** | 0.95[0.66, 1.36] | 0.7670 | 0.40[0.23, 0.70] | 0.0014** |
| Mean corpuscular volume, fL | 1.02[1.01, 1.03] | 0.0001*** | 1.01[0.99, 1.03] | 0.5120 | 1.02[0.99, 1.05] | 0.1190 |
| Basophil, x10<sup>9</sup>/L | 1.18[0.66, 2.12] | 0.5820 | 1.54[0.92, 2.59] | 0.1010 | 1.51[0.72, 3.16] | 0.2770 |
| Eosinophil, x10<sup>9</sup>/L | 1.00[0.76, 1.30] | 0.9830 | 1.20[0.97, 1.48] | 0.0973. | 1.18[0.89, 1.55] | 0.2460 |
| Lymphocyte, x10<sup>9</sup>/L | 0.59[0.53, 0.65] | <0.0001*** | 0.86[0.70, 1.07] | 0.1710 | 0.82[0.64, 1.04] | 0.1020 |
| Monocyte, x10<sup>9</sup>/L | 1.68[1.27, 2.22] | 0.0002*** | 2.40[1.36, 4.22] | 0.0025** | 4.35[2.71, 6.98] | <0.0001*** |
| Neutrophil, x10<sup>9</sup>/L | 1.06[1.05, 1.08] | <0.0001*** | 1.07[1.04, 1.10] | <0.0001*** | 1.06[1.02, 1.10] | 0.0016** |
| White blood cells, x10<sup>9</sup>/L | 1.024[1.018, 1.030] | <0.0001*** | 1.03[1.03, 1.04] | <0.0001*** | 1.02[1.00, 1.04] | 0.0256* |
| Mean cell haemoglobin, pg | 1.02[1.00, 1.04] | 0.0953. | 0.99[0.94, 1.04] | 0.5970 | 1.003[0.947, 1.063] | 0.9070 |
| Myelocyte, x10<sup>9</sup>/L | 0.58[0.14, 2.32] | 0.4390 | 0.93[0.24, 3.57] | 0.9120 | 1.000[0.815, 1.227] | 0.9990 |
| Platelet, x10<sup>9</sup>/L | 1.00[1.00, 1.00] | 0.0016** | 1.002[1.000, 1.004] | 0.0525. | 0.995[0.992, 0.998] | 0.0002*** |
| Reticulocyte, x10<sup>9</sup>/L | 1.00[0.98, 1.01] | 0.7560 | 1.000[0.980, 1.021] | 0.9740 | 0.96[0.91, 1.02] | 0.1470 |
| Red blood cells, x10<sup>12</sup>/L | 0.55[0.50, 0.62] | <0.0001*** | 0.67[0.52, 0.86] | 0.0015** | 0.83[0.63, 1.09] | 0.1800 |
| Hematocrit, L/L | 0.00[0.00, 0.01] | <0.0001*** | 0.00[0.00, 0.10] | 0.0011** | 0.26[0.00, 14.31] | 0.5140 |
| Potassium, mmol/L | 1.12[0.99, 1.27] | 0.0719. | 0.95[0.70, 1.29] | 0.7480 | 1.15[0.84, 1.57] | 0.3920 |
| Urate, mmol/L | 11.6[3.3, 41.2] | 0.0002*** | 183.0[15.2, 2205.0] | <0.0001*** | 22.17[2.13, 230.50] | 0.0095** |
| Albumin, g/L | 0.87[0.86, 0.88] | <0.0001*** | 0.86[0.84, 0.89] | <0.0001*** | 0.96[0.92, 1.00] | 0.0409* |
| Sodium, mmol/L | 0.94[0.92, 0.96] | <0.0001*** | 0.89[0.86, 0.93] | <0.0001*** | 1.02[0.97, 1.07] | 0.5570 |
| Urea, mmol/L | 1.07[1.06, 1.09] | <0.0001*** | 1.06[1.02, 1.09] | 0.0008*** | 1.06[1.03, 1.09] | 0.0003*** |
| Protein, g/L | 0.95[0.94, 0.96] | <0.0001*** | 0.98[0.95, 1.00] | 0.0780. | 0.98[0.95, 1.01] | 0.2140 |
| Creatinine, umol/L | 1.002[1.002, 1.003] | <0.0001*** | 1.002[1.001, 1.003] | 0.0006*** | 1.002[1.001, 1.003] | 0.0054** |
| Alkaline phosphatase, U/L | 1.01[1.01, 1.01] | <0.0001*** | 1.006[1.004, 1.008] | <0.0001*** | 0.999[0.993, 1.005] | 0.7260 |
| Aspartate transaminase, U/L | 1.004[1.001, 1.006] | 0.0059** | 1.004[0.999, 1.009] | 0.1560 | 0.998[0.983, 1.014] | 0.8170 |
| Alanine transaminase, U/L | 0.997[0.994, 1.001] | 0.1130 | 0.996[0.988, 1.004] | 0.3510 | 0.988[0.977, 0.999] | 0.0389* |
| Bilirubin, umol/L | 1.012[1.007, 1.018] | <0.0001*** | 1.02[1.01, 1.03] | <0.0001*** | 1.02[1.01, 1.03] | <0.0001*** |
| Triglyceride, mmol/L | 0.94[0.89, 0.99] | 0.0120* | 0.96[0.85, 1.07] | 0.4200 | 0.87[0.75, 1.00] | 0.0543 |
| Total cholesterol, mmol/L | 0.95[0.91, 1.00] | 0.0559. | 0.96[0.86, 1.08] | 0.5170 | 0.87[0.77, 0.98] | 0.0195* |
| LDL, mmol/L | 1.07[0.99, 1.15] | 0.0816. | 0.91[0.76, 1.09] | 0.3180 | 0.77[0.63, 0.94] | 0.0098** |
| HDL, mmol/L | 0.75[0.62, 0.92] | 0.0053** | 0.27[0.16, 0.46] | <0.0001*** | 0.27[0.16, 0.47] | <0.0001*** |
| Fasting blood glucose, mmol/L | 1.03[1.02, 1.04] | <0.0001*** | 1.04[1.02, 1.07] | 0.0003*** | 0.96[0.92, 1.01] | 0.0957 |
| HbA1c, g/dL | 0.99[0.98, 1.00] | 0.0537. | 0.97[0.94, 0.99] | 0.0113* | 0.98[0.95, 1.01] | 0.1480 |

HF: heart failure; IHD: ischemic heart disease; COPD: chronic obstructive pulmonary disease; LDL: low-density lipoprotein cholesterol; HDL: high-density lipoprotein cholesterol.

**Table 4.** Cox regression models for all-cause mortality, cardiovascular mortality and new onset stroke after propensity score matching (1:2). * for p≤ 0.05, ** for p ≤ 0.01, *** for p ≤ 0.001

| Characteristics | All-cause mortality<br>HR [95% CI] | P value | Cardiovascular mortality<br>HR [95% CI] | P value | New onset stroke<br>HR [95% CI] | P value |
| --- | --- | --- | --- | --- | --- | --- |
| Male gender | 1.27[1.14, 1.41] | <0.0001*** | 1.16[0.91, 1.47] | 0.2310 | 1.07[0.87, 1.31] | 0.5440 |
| Baseline age, year | 1.08[1.07, 1.08] | <0.0001*** | 1.08[1.06, 1.09] | <0.0001*** | 1.09[1.08, 1.10] | <0.0001*** |
| <50 | 0.25[0.20, 0.31] | <0.0001*** | 0.30[0.18, 0.49] | <0.0001*** | 0.16[0.09, 0.28] | <0.0001*** |
| [50-60] | 0.50[0.44, 0.57] | <0.0001*** | 0.54[0.41, 0.72] | <0.0001*** | 0.34[0.25, 0.45] | <0.0001*** |
| [60-70] | 0.77[0.69, 0.86] | <0.0001*** | 0.70[0.54, 0.90] | 0.0063** | 1.06[0.86, 1.31] | 0.5590 |
| [70-80] | 2.56[2.30, 2.85] | <0.0001*** | 2.42[1.88, 3.11] | <0.0001*** | 3.33[2.70, 4.10] | <0.0001*** |
| >80 | 6.07[5.35, 6.89] | <0.0001*** | 6.23[4.68, 8.31] | <0.0001*** | 4.41[3.30, 5.90] | <0.0001*** |
| Cardiovascular | 5.1[4.6, 5.6] | <0.0001*** | 16.97[12.8, 22.5] | <0.0001*** | 3.11[2.53, 3.80] | <0.0001*** |
| Respiratory | 6.51[5.89, 7.19] | <0.0001*** | 6.23[4.95, 7.84] | <0.0001*** | 1.95[1.56, 2.43] | <0.0001*** |
| Renal | 3.41[3.03, 3.84] | <0.0001*** | 3.59[2.74, 4.70] | <0.0001*** | 1.82[1.36, 2.44] | 0.0001*** |
| Endocrine | 3.86[3.12, 4.77] | <0.0001*** | 0.65[0.21, 2.04] | 0.4650 | 2.59[1.55, 4.35] | 0.0003*** |
| Hypertension | 1.98[1.79, 2.18] | <0.0001*** | 1.80[1.43, 2.27] | <0.0001*** | 6.46[5.15, 8.10] | <0.0001*** |
| Gastrointestinal | 2.57[2.32, 2.86] | <0.0001*** | 1.03[0.76, 1.40] | 0.8520 | 1.42[1.11, 1.82] | 0.0046** |
| HF | 7.18[5.87, 8.79] | <0.0001*** | 8.90[5.86, 13.51] | <0.0001*** | 0.87[0.28, 2.72] | 0.8140 |
| IHD | 2.81[2.50, 3.15] | <0.0001*** | 5.92[4.69, 7.46] | <0.0001*** | 2.45[1.92, 3.13] | <0.0001*** |
| Dementia | 8.20[3.68, 18.28] | <0.0001*** | 6.84[0.96, 48.73] | 0.0550. | - | - |
| COPD | 4.91[0.69, 34.91] | 0.1120 | - | - | - | - |
| Cancer | 5.13[4.20, 6.27] | <0.0001*** | 1.24[0.51, 3.00] | 0.6360 | 2.35[1.29, 4.28] | 0.0052** |
| Charlson score | 1.79[1.74, 1.83] | <0.0001*** | 1.75[1.66, 1.85] | <0.0001*** | 1.69[1.60, 1.79] | <0.0001*** |
| SGLT2I | 0.64[0.56, 0.74] | <0.0001*** | 0.74[0.55, 1.00] | 0.0470* | 0.46[0.33, 0.64] | <0.0001*** |
| DPP4I | 1.55[1.35, 1.78] | <0.0001*** | 1.36[1.00, 1.83] | 0.0470* | 2.19[1.57, 3.06] | <0.0001*** |
| Gliclazide | 1.34[1.22, 1.49] | <0.0001*** | 1.82[1.43, 2.32] | <0.0001*** | 1.77[1.43, 2.18] | <0.0001*** |
| Glimepiride | 0.81[0.69, 0.95] | 0.0095** | 0.82[0.57, 1.19] | 0.2990 | 0.81[0.58, 1.11] | 0.1860 |
| Metformin | 0.98[0.89, 1.09] | 0.7590 | 1.05[0.82, 1.34] | 0.6840 | 1.45[1.15, 1.83] | 0.0016** |
| Sulphonylurea | 0.76[0.64, 0.91] | 0.0031** | 0.70[0.45, 1.08] | 0.1080 | 0.85[0.60, 1.19] | 0.3360 |
| Insulin | 1.06[0.95, 1.17] | 0.3190 | 1.12[0.88, 1.43] | 0.3410 | 0.84[0.67, 1.04] | 0.1110 |
| Thiozolidinedone | 0.51[0.38, 0.67] | <0.0001*** | 0.58[0.31, 1.10] | 0.0951. | 1.56[1.10, 2.20] | 0.0116* |
| Meglitinide | 0.79[0.66, 0.95] | 0.0104* | 0.71[0.45, 1.10] | 0.1280 | 0.79[0.55, 1.13] | 0.1960 |
| Glucagon-like peptide-1 agonist | 0.60[0.39, 0.93] | 0.0221* | 0.17[0.02, 1.19] | 0.0736. | 3.19[2.15, 4.72] | <0.0001*** |
| Acarbose | 1.66[1.08, 2.56] | 0.0206* | 0.85[0.21, 3.40] | 0.8130 | 1.61[0.67, 3.90] | 0.2880 |
| Diuretics for hypertension | 0.58[0.33, 1.00] | 0.0483* | 0.25[0.03, 1.75] | 0.1610 | 0.61[0.19, 1.88] | 0.3860 |
| Diuretics for HF | 1.98[1.47, 2.68] | <0.0001*** | 5.17[3.28, 8.15] | <0.0001*** | 0.21[0.03, 1.47] | 0.1150 |
| Anticoagulants | 1.37[0.98, 1.92] | 0.0644. | 4.13[2.56, 6.66] | <0.0001*** | 0.84[0.35, 2.03] | 0.7000 |
| Antiplatelets | 0.94[0.76, 1.15] | 0.513 | 1.62[1.11, 2.36] | 0.0131* | 0.66[0.41, 1.06] | 0.0848. |
| Lipid-lowering drugs | 0.60[0.50, 0.71] | <0.0001*** | 0.82[0.57, 1.17] | 0.2670 | 0.61[0.42, 0.89] | 0.0091** |
| Mean corpuscular volume, fL | 1.03[1.02, 1.04] | <0.0001*** | 1.01[0.99, 1.03] | 0.2490 | 1.04[1.02, 1.06] | 0.0013** |
| Basophil, x10<sup>9</sup>/L | 1.01[0.40, 2.54] | 0.9790 | 1.45[0.79, 2.67] | 0.2270 | 1.49[0.70, 3.15] | 0.3000 |
| Eosinophil, x10<sup>9</sup>/L | 1.05[0.85, 1.30] | 0.6590 | 1.18[0.95, 1.47] | 0.1420 | 1.25[1.02, 1.52] | 0.0302* |
| Lymphocyte, x10<sup>9</sup>/L | 0.54[0.49, 0.60] | <0.0001*** | 0.70[0.57, 0.86] | 0.0008*** | 1.12[1.04, 1.20] | 0.0017** |
| Monocyte, x10<sup>9</sup>/L | 1.92[1.47, 2.50] | <0.0001*** | 2.42[1.42, 4.12] | 0.0012** | 4.85[3.18, 7.40] | <0.0001*** |
| Neutrophil, x10<sup>9</sup>/L | 1.06[1.05, 1.08] | <0.0001*** | 1.06[1.03, 1.09] | <0.0001*** | 1.07[1.03, 1.10] | 0.0001*** |
| White blood cells, x10<sup>9</sup>/L | 1.023[1.016, 1.029] | <0.0001*** | 1.03[1.02, 1.04] | <0.0001*** | 1.04[1.03, 1.04] | <0.0001*** |
| Mean cell haemoglobin, pg | 1.03[1.00, 1.05] | 0.0217* | 0.99[0.95, 1.04] | 0.7220 | 1.08[1.02, 1.14] | 0.0124* |
| Myelocyte, x10<sup>9</sup>/L | 0.57[0.13, 2.43] | 0.4470 | 0.93[0.28, 3.14] | 0.9120 | 0.95[0.36, 2.54] | 0.9200 |
| Platelet, x10<sup>9</sup>/L | 0.998[0.997, 0.999] | <0.0001*** | 1.000[0.998, 1.002] | 0.8600 | 1.00[1.00, 1.00] | 0.4410 |
| Reticulocyte, x10<sup>9</sup>/L | 0.996[0.981, 1.010] | 0.5670 | 1.002[0.980, 1.025] | 0.8430 | - | - |
| Red blood cells, x10<sup>12</sup>/L | 0.52[0.47, 0.57] | <0.0001*** | 0.62[0.49, 0.78] | <0.0001*** | 0.68[0.53, 0.85] | 0.0011** |
| Hematocrit, L/L | 0.00[0.00, 0.00] | <0.0001*** | 0.00[0.00, 0.02] | <0.0001*** | 0.03[0.00, 0.68] | 0.0285* |
| Potassium, mmol/L | 1.11[0.98, 1.25] | 0.1100 | 0.95[0.71, 1.26] | 0.7060 | 1.19[0.93, 1.52] | 0.1650 |
| Urate, mmol/L | 14.06[4.25, 46.51] | <0.0001*** | 730.50[82.49, 6470.00] | <0.0001*** | 0.13[0.01, 1.32] | 0.0835. |
| Albumin, g/L | 0.87[0.86, 0.88] | <0.0001*** | 0.87[0.85, 0.89] | <0.0001*** | 0.91[0.89, 0.94] | <0.0001*** |
| Sodium, mmol/L | 0.94[0.93, 0.96] | <0.0001*** | 0.89[0.86, 0.92] | <0.0001*** | 1.01[0.97, 1.05] | 0.5820 |
| Urea, mmol/L | 1.08[1.07, 1.09] | <0.0001*** | 1.07[1.04, 1.10] | <0.0001*** | 1.03[1.00, 1.06] | 0.0654. |
| Protein, g/L | 0.96[0.95, 0.97] | <0.0001*** | 0.99[0.96, 1.01] | 0.3220 | 1.03[1.00, 1.05] | 0.0218* |
| Creatinine, umol/L | 1.002[1.002, 1.003] | <0.0001*** | 1.002[1.002, 1.003] | <0.0001*** | 1.001[1.000, 1.003] | 0.0119* |
| Alkaline phosphatase, U/L | 1.006[1.005, 1.007] | <0.0001*** | 1.006[1.004, 1.008] | <0.0001*** | 1.002[0.999, 1.005] | 0.2230 |
| Aspartate transaminase, U/L | 1.004[1.001, 1.006] | 0.0066** | 1.003[0.996, 1.009] | 0.4200 | 0.94[0.91, 0.98] | 0.0024** |
| Alanine transaminase, U/L | 0.999[0.996, 1.002] | 0.3800 | 1.000[0.994, 1.006] | 0.8810 | 0.98[0.96, 0.99] | <0.0001*** |
| Bilirubin, umol/L | 1.014[1.009, 1.018] | <0.0001*** | 1.016[1.008, 1.024] | 0.0001*** | 1.02[1.01, 1.02] | 0.0009*** |
| Triglyceride, mmol/L | 0.91[0.87, 0.96] | 0.0005*** | 0.95[0.86, 1.06] | 0.3630 | 0.89[0.80, 0.98] | 0.0243* |
| Total cholesterol, mmol/L | 0.93[0.88, 0.97] | 0.0013** | 0.97[0.87, 1.08] | 0.5270 | 0.84[0.77, 0.92] | 0.0001*** |
| LDL, mmol/L | 1.03[0.96, 1.11] | 0.4630 | 0.93[0.79, 1.11] | 0.4250 | 0.85[0.73, 0.99] | 0.0342* |
| HDL, mmol/L | 0.76[0.63, 0.92] | 0.0059** | 0.33[0.20, 0.55] | <0.0001*** | 0.87[0.61, 1.25] | 0.4570 |
| Fasting blood glucose, mmol/L | 1.03[1.01, 1.04] | <0.0001*** | 1.04[1.02, 1.06] | 0.0012** | 1.02[0.99, 1.04] | 0.2220 |
| HbA1c, g/dL | 0.98[0.97, 0.99] | 0.0046** | 0.97[0.94, 0.99] | 0.0089** | 1.00[0.97, 1.03] | 0.9100 |

HF: heart failure; IHD: ischemic heart disease; COPD: chronic obstructive pulmonary disease; LDL: low-density lipoprotein cholesterol; HDL: high-density lipoprotein cholesterol.

**Table 5.** Multivariate cox analysis for all-cause mortality, cardiovascular mortality and new onset AF after propensity score matching (1:2). * for p≤ 0.05, ** for p ≤ 0.01, *** for p ≤ 0.001

| Characteristics |  | All-cause mortality<br>HR [95% CI] | P value | Cardiovascular mortality<br>HR [95% CI] | P value | New onset AF<br>HR [95% CI] | P value |
| --- | --- | --- | --- | --- | --- | --- | --- |
| Model 1 | SGLT2I (DPP4I as reference) | 0.69[0.60, 0.79] | <0.0001*** | 0.79[0.58, 1.09] | 0.1510 | 0.43[0.28, 0.66] | 0.0001*** |
|  | DPP4I (SGLT2I as reference) | 1.45[1.27, 1.67] | <0.0001*** | 1.26[0.92, 1.72] | 0.1510 | 2.34[1.51, 3.60] | 0.0001*** |
| Model 2 | SGLT2I (DPP4I as reference) | 0.76[0.66, 0.87] | <0.0001*** | 0.82[0.60, 1.12] | 0.2200 | 0.58[0.38, 0.91] | 0.0161* |
|  | DPP4I (SGLT2I as reference) | 1.32[1.15, 1.5] | <0.0001*** | 1.22[0.89, 1.66] | 0.2200 | 1.72[1.11, 2.66] | 0.0161* |
| Model 3 | SGLT2I (DPP4I as reference) | 0.72[0.63,0.82] | <0.0001*** | 0.78[0.57, 1.07] | 0.1220 | 0.58[0.37, 0.90] | 0.0143* |
|  | DPP4I (SGLT2I as reference) | 1.39[1.21,1.60] | <0.0001*** | 1.28[0.94, 1.75] | 0.1220 | 1.73[1.11, 2.69] | 0.0143* |
| Model 4 | SGLT2I (DPP4I as reference) | 0.68[0.57, 0.79] | <0.0001*** | 0.75[0.54, 1.03] | 0.7810 | 0.57[0.36, 0.88] | 0.0132* |
|  | DPP4I (SGLT2I as reference) | 1.36[1.18, 1.54] | <0.0001*** | 1.23[0.88, 1.66] | 0.7810 | 1.71[1.08, 2.63] | 0.0132* |

IR: incidence rate.
Model 1 adjusted for none.
Model 2 adjusted for age and gender.
Model 3 adjusted for age, gender, cardiovascular, respiratory, renal, endocrine, hypertension, gastrointestinal, HF, IHD, dementia, COPD, cancer, and Charlson score.
Model 4 adjusted for gliclazide, glimepiride, metformin, sulphonylurea, insulin, thiazolidinedione, meglitinide, glucagon-like peptide-1 agonist, acarbose, diuretics for hypertension, diuretics for HF, anticoagulants, antiplatelets, lipid-lowering drugs in model 3.

**Table 6.**
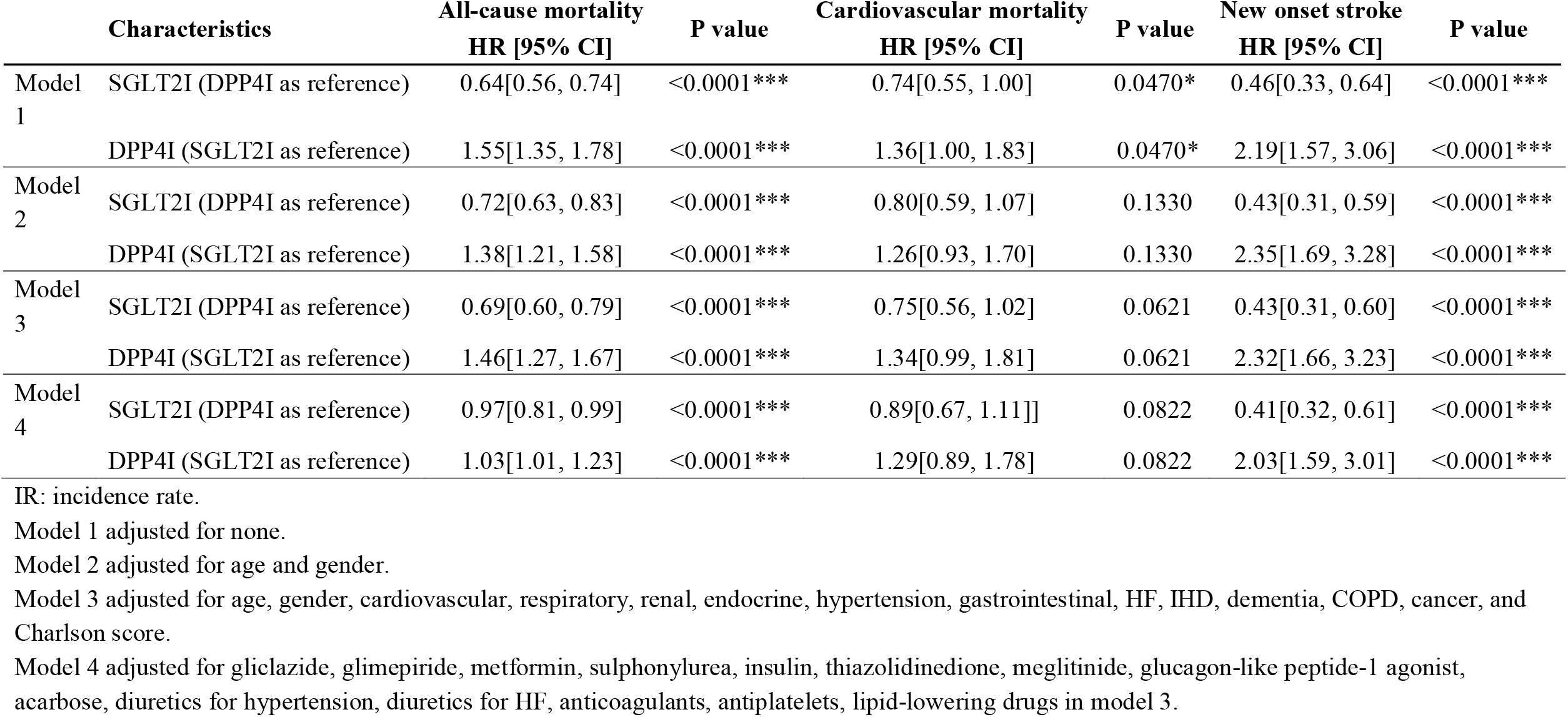
Multivariate cox analysis for all-cause mortality, cardiovascular mortality and new onset stroke after propensity score matching (1:2). * for p≤ 0.05, ** for p ≤ 0.01, *** for p ≤ 0.001

Those with new onset stroke presentation had lower levels of reticulocyte (mean [±SD]: 60.81±23.54; max: 111.0 x10^9/L v.s. mean [±SD]:69.34±29.19; max: 168.0 x10^9/L, SMD=0.32), urate (mean [±SD]: 0.37±0.1; max: 0.91 mmol/L v.s. mean [±SD]:0.41±0.12; max: 0.97 mmol/L, SMD=0.31), urea (mean [±SD]: 5.59±1.78; max: 44.6 mmol/L v.s. mean [±SD]:6.36±2.98; max: 43.21 mmol/L, SMD=0.31), creatinine (mean [±SD]: 77.49±21.66; max: 757.0 umol/L v.s. mean [±SD]:91.38±57.16; max: 1294.0 umol/L, SMD=0.32), in addition to higher levels of red blood cells (mean [±SD]: 4.77±0.59; max: 9.18 x10^12/L v.s. mean [±SD]:4.61±0.65; max: 7.86 x10^12/L, SMD=0.26), HCT (mean [±SD]: 0.41±0.04; max: 0.61 L/L v.s. mean [±SD]:0.4±0.05; max: 0.59 L/L, SMD=0.33) and ALT (mean [±SD]: 34.04±30.18; max: 738.0 L/L v.s. mean [±SD]:28.03±23.85; max: 605.0 L/L, SMD=0.22).

## Discussion

There are several major findings for the present study: 1) SGLT2I users have a lower risk for new-onset stroke and AF in comparison to DPP4I users; 2) SGLT2I users have a lower risk for all-cause mortality, cardiovascular mortality, new-onset stroke and AF; 3) DPP4I use increases the risk of all-cause mortality, cardiovascular mortality, new-onset stroke and AF.

To our knowledge, this is the first study with a head-to-head comparison of new-onset stroke and AF risk between SGLT2I and DPP4I users. Whilst the superior effects of SGLT2I against DPP4I in the lower of all-cause and cardiovascular mortality have been reported, few studies have compared the effects of the two drug classes in their effects on stroke and AF specifically.(7) An existing meta-analysis, which compared the stroke risk amongst SGLT2I and DPP4I users indirectly, reported that the effect of both drug classes was neutral. (15) However, the meta-analysis had a smaller sample size, and confounders were not adjusted for. Recently, a multinational observational study involving 13 countries demonstrated significantly lower all-cause mortality and stroke risk in SGLT2I users in comparison to DPP4I users, which is supportive of the present findings. (8)

On the other hand, the effect of SGLT2I on AF is more controversial. The risk of AF was numerically lower in all, and statistically lower in some clinical trials comparing SGLT2I users and controls. (16-19) A recent meta-analysis of 16 randomized controlled trials demonstrated a significant reduction of AF and atrial flutter amongst SGLT2I and placebo users. (20) Although findings from the Cardiovascular mortality and morbidity in patients with type 2 diabetes following initiation of sodium-glucose co-transporter-2 inhibitors versus other glucose-lowering drugs (CVD-REAL Nordic) study demonstrated an insignificant difference in stroke and atrial fibrillation between dapagliflozin and DPP4I users, their analysis did not adjust for clinical or biochemical confounders, nor a history of the two diseases respectively. (21) Animal studies have also demonstrated that SGLT2I can prevent atrial remodeling, which is an important pathogenic mechanism of AF. (22) Thus, the findings from the present study, which is based on more stringent patient selection and confounder adjustments, and better supported by the large-scale trials on SGLT2I are likely a better reflection of the clinical circumstances.

Existing reports on the cardiovascular effects of DPP4I remains controversial. Insignificant differences in cardiovascular mortality, myocardial infarction and ischemic stroke between DPP4I users and controls were reported by both the Cardiovascular Outcomes Study of Alogliptin in Patients With Type 2 Diabetes and Acute Coronary Syndrome (EXAMINE) and the Saxagliptin Assessment of Vascular Outcomes Recorded in Patients With Diabetes Mellitus–Thrombolysis in Myocardial Infarction 53 (SAVOR-TIMI53) trials. (23, 24) Hospitalization for acute HF increased in the SAVOR-TIMI53 trial, though the results were not replicated by other trials. (23-25) An insignificant elevation in risk of HF-related hospitalization was reported in a meta-analysis summarizing these clinical trials, with substantial heterogeneity across trials using different DPP4Is, suggesting unique features of specific DPP4I may have individual cardiovascular effects. (26)

### Limitations

Several limitations should be noted for the present study. First of all, given its observational nature, there is inherent information bias due to under-coding, coding errors and missing data. Secondly, residual confounding may be present despite robust propensity-matching, particularly with the unavailability of information on lifestyle cardiovascular risk factors. Patients’ drug exposure duration has not been controlled, which may affect their risk against the study outcomes. Additionally, the occurrence of AF out of the hospital is not accounted for, though non-sustained AF that does not result in hospital admission can be considered as subclinical.

## Conclusions

## Data Availability

Data available upon request.

## Conflicts of Interest

None.

## Notes

### Competing Interest Statement

The authors have declared no competing interest.

### Author Declarations

The study was approved by The Joint Chinese University of Hong Kong - New Territories East Cluster Clinical Research Ethics Committee and Institutional Review Board of the University of Hong Kong/Hospital Authority Hong Kong West Cluster.

