## Supplementary Appendix for "Comparative effects of sodium glucose cotransporter 2 (SGLT2) inhibitors and dipeptidyl peptidase-4 (DPP4) inhibitors on new-onset atrial fibrillation and stroke outcomes"

**Supplementary Table 1. ICD9 codes for comorbidities and ICD10 codes for outcomes**

Cardiovascular	427.31	423.9	427.5	428	785.1	413.9	427.89	410.71	785	794.31	428.9	427.81	427.69	427.32	427	426.4	426.11	426.1	425.4	424.9	424	421	398.9	787.01	151	414.9	V72.81	410.01	V43.3	164.1	402.9	785.51	426	414.8	428.1	309	402.91	404.93	746.87	V81.0	414	648.63	296.53	300.4	411	426.13	528.3	996.2	416.9	429.3	787.1	785.9	427.2	786.51	414.1	398.91	426.12	391.9	404.11	416	416.8	746.89	745.8	746	746.86	746.9	745.9	648.5	E878.0	648.51	V53.31	404.9	746.7	404.91	404.92	404.01	648.53	V71.7	398	397.9	E879.0	410	410.9	745.6	648.61	397	429.4	443.9	412	423	V45.00	972	785.2	427.1	745.69	431	996.63

Respiratory	786.09	518.81	780.53	137	E912	465.9	79.6	519.8	780.59	799.1	780.57	518.82	480.1	786.3	997.3	165.9	519.9	648.91	162.9	162.3	197	162.5	162.4	486	518.89	496	162.8	415.1	V10.11	518	11.96	482.1	507	513	11.9	511.8	511.1	11.94	516.8	793.1	482.4	515	11.93	482	482.83	518.4	482.3	482.2	502	235.7	934.8	516.9	136.3	38.49	506	112.4	487	481	117.9	38.2	11.95	79.89	518.1	480.9	505	495.9	518.3	11.23	416.8	397.1	117.3	483	508	998.81	416	514	861.21	480.8	648.93	11.2	492.8	484.6	78.5	484.1	516.3	416.9	415	429.89	747.49	745	417	770.7	427.5	746.02	573.8	642.9	747.3	770.3	779.8	424.3	417.8	745.4	V12.6	478	748.5	996.84	748.6	V42.1	11.05	162	747.42	517.2

Renal	198.7	189	585.9	V56.0	189.1	584.9	593.9	189.8	239.5	583.81	591	V10.52	250.4	255.4	590.8	586	585.1	239.7	788	996.39	588.9	592	593.2	572.4	194	255	453.3	198	590.1	223	250.41	581.9	227	593.5	583.89	593.89	404.93	255.5	788.9	250.43	405.92	753.12	E879.1	V42.0	580.89	403.9	V59.4	580.9	794.4	584.8	584.5	255.9	441.4	V58.49	404.11	585	759.1	753.11	753.15	274	588.8	403.91	404.9	404.91	404.92	403	404.01	779.8	996.81	753.8	753	589.1	582.9	753.1	753.3	E878.0	753.17	583.9	589	866	404	580	581	582	583	584	590

Endocrine	202.8	200.1	200.12	201.9	204	202.88	200.18	196	204.01	785.6	200.11	200.13	202.85	202.81	196.9	202.82	202	202.87	V10.79	202.84	202.01	196.8	457.1	12.1	196.5	196.2	238.7	196.1	200.14	457.2	V10.61	457	289.3	245.2	457.9	202.93	202.97	757	V10.71	288.8	204.1	202.83	V77.9	237.4	239.7	198.89	623.5	259.9	200.2

Hypertension	401.9	250.82	790.6	796.2	402.9	250.83	405.99	642.93	642.01	642.91	E942.6	405.09	403.9	437.2	401	401.1	642.33	348.2	779.8	365.04	572.3	416	405.91	416.8	642.3

Gastrointestinal	153.3	154.1	153.9	569.89	154	153.1	578.9	560.9	569.3	537.89	558.9	562.1	153.6	239	532.3	532.7	535.6	38.42	8.45	153.2	569.49	79.89	532.9	V58.11	569	41.4	152.1	V10.05	787.8	197.4	535.5	V10.06	9	569.83	569.6	153.4	537.3	41.04	569.84	569.81	8.8	535	532	V45.89	V12.72	532.4	V10.09	560.81	235.2	38.49	557.9	569.41	997.4	14.8	787.99	8.46	569.82	537.9	560.1	211.3	556.9	562	536.9	566	V71.9	564.3	V44.4	564.8	562.11	211.2	9.1	8.47	8.5	532.1	535.61	560	565.1	619.1	152.9	568	569.43	152	8.61	596.1	151.4	151.9	151.5	151.8	151.1	456.8	531.7	535.4	531.3	V15.2	211.1	531.9	V10.04	531	531.4	535.1	151.3	230.2	151.6	536.3	535.51	531.1	531.5	537.84	535.01	530.7	535.2	537.6	202.83	558	569.85	153	555.1	562.13	562.12	V76.49	560.2	230.4

Atrial fibrillation	427.31	429.4

Transient ischemic attack /ischemic stroke 435	435	435.1	435.2	435.3	435.8	435.9	433.81	433.91	434	436	437	437.1	433.31	433.01	434	434	434	434.01	434.1	434.1	434.11	434.9	434.9	434.91	436	437	437	437.1	437.2	437.3	437.4	437.5	437.6	437.7	437.8	437.9

Ischemic heart disease	410.01	410.02	410.1	410.11	410.12	410.2	410.21	410.22	410.3	410.31	410.32	410.4	410.41	410.42	410.5	410.51	410.52	410.6	410.61	410.62	410.7	410.71	410.72	410.8	410.81	410.82	410.9	410.91	410.92	411	411.1	411.8	411.81	411.89	413	413.1	413.9	414	414.01	414.02	414.03	414.04	414.05	414.06	414.07	414.1	414.11	414.12	414.19	414.2	414.3	414.4	414.8	414.9	410	412

Sudden cardiac death	427.01	427.4	427.5	410	410.01	410.02	410.1	410.11	410.12	410.2	410.21	410.22	410.3	410.31	410.32	410.4	410.41	410.42	410.5	410.51	410.52	410.6	410.61	410.62	410.7	410.71	410.72	410.8	410.81	410.82	410.9	410.91	410.92

Dementia 331.82	290	290.1	290.11	290.12	290.13	290.2	290.21	290.3	290.4	290.41	290.42	290.43	290.8	290.9	294.2	294.1	294.11	294.21	332	46.1	333.4	340	42	331	331.19	294.29

Chronic Obstructive PD	490	491	492	493	494	495	496	491.1	491.2	491.21	491.22	491.8	491.9	492.8	493.01	493.02	493.1	493.11	493.12	493.2	493.21	493.22	493.8	493.81	493.82	493.9	493.91	493.92	494.1	495.1	495.2	495.3	495.4	495.5	495.6	495.7	495.8	495.9

Cancer 140-239

Heart failure	57.9	79.99	198.5	211.4	225.4	288.3	303.02	389.11	398.9	402.91	416.9	427.5	428	428.1	428.9	429.4	478.31	611.79	617.3	621.8	711.09	727.64	784.4	891.1	916.9	V22.1	428.99

Cardiovascular mortality

I00-I09, I11, I13, I20-I51

**Supplementary Table 2. Baseline characteristics of overall patients and patients with/without new onset AF.**

* for SMD$\geq$0.2; # indicates the difference between patients with/without new onset AF.

| **Characteristics** | **Overall (N=49108)**  **Mean(SD);Max;N or Count(%)** | **New AF (N=842)**  **Mean(SD);Max;N or Count(%)** | **No new AF (N=48266)**  **Mean(SD);Max;N or Count(%)** | **SMD^#^** |
| --- | --- | --- | --- | --- |
| Male gender | 27171(55.32%) | 457(54.27%) | 26714(55.34%) | 0.02 |
| Baseline age, year | 66.48(12.89);104.55;n=49108 | 77.5(10.0);99.7;n=842 | 66.3(12.8);104.5;n=48266 | 0.97* |
| <50 | 4623(9.41%) | 7(0.83%) | 4616(9.56%) | 0.40* |
| [50-60] | 10538(21.45%) | 46(5.46%) | 10492(21.73%) | 0.49* |
| [60-70] | 15086(30.72%) | 129(15.32%) | 14957(30.98%) | 0.38* |
| [70-80] | 10664(21.71%) | 270(32.06%) | 10394(21.53%) | 0.24* |
| >80 | 8197(16.69%) | 390(46.31%) | 7807(16.17%) | 0.69* |
| Cardiovascular | 11705(23.83%) | 842(100.00%) | 10863(22.50%) | 2.62* |
| Respiratory | 12143(24.72%) | 499(59.26%) | 11644(24.12%) | 0.76* |
| Renal | 11382(23.17%) | 428(50.83%) | 10954(22.69%) | 0.61* |
| Endocrine | 1017(2.07%) | 29(3.44%) | 988(2.04%) | 0.09 |
| Hypertension | 26119(53.18%) | 643(76.36%) | 25476(52.78%) | 0.51* |
| Gastrointestinal | 13126(26.72%) | 398(47.26%) | 12728(26.37%) | 0.44* |
| HF | 688(1.40%) | 30(3.56%) | 658(1.36%) | 0.14 |
| IHD | 4890(9.95%) | 130(15.43%) | 4760(9.86%) | 0.17 |
| Dementia | 550(1.11%) | 12(1.42%) | 538(1.11%) | 0.03 |
| COPD | 10(0.02%) | 0(0.00%) | 10(0.02%) | 0.02 |
| Cancer | 1419(2.88%) | 43(5.10%) | 1376(2.85%) | 0.12 |
| Charlson score | 2.55(1.74);16.0;n=49108 | 3.9(1.7);14.0;n=842 | 2.5(1.7);16.0;n=48266 | 0.82* |
| SGLT2I | 12526(25.50%) | 23(2.73%) | 12503(25.90%) | 0.70* |
| DPP4I | 36582(74.49%) | 819(97.26%) | 35763(74.09%) | 0.70* |
| Gliclazide | 33307(67.82%) | 610(72.44%) | 32697(67.74%) | 0.10 |
| Glimepiride | 6338(12.90%) | 105(12.47%) | 6233(12.91%) | 0.01 |
| Metformin | 34167(69.57%) | 568(67.45%) | 33599(69.61%) | 0.05 |
| Sulphonylurea | 4857(9.89%) | 97(11.52%) | 4760(9.86%) | 0.05 |
| Insulin | 10857(22.10%) | 188(22.32%) | 10669(22.10%) | 0.01 |
| Thiozolidinedone | 860(1.75%) | 5(0.59%) | 855(1.77%) | 0.11 |
| Meglitinide | 4371(8.90%) | 90(10.68%) | 4281(8.86%) | 0.06 |
| Glucagon-like peptide-1 agonist | 193(0.39%) | 0(0.00%) | 193(0.39%) | 0.09 |
| Acarbose | 389(0.79%) | 21(2.49%) | 368(0.76%) | 0.14 |
| Diuretics for hypertension | 643(1.30%) | 3(0.35%) | 640(1.32%) | 0.11 |
| Diuretics for HF | 675(1.37%) | 1(0.11%) | 674(1.39%) | 0.15 |
| Lipid-lowering drugs | 7214(14.69%) | 13(1.54%) | 7201(14.91%) | 0.50* |
| Anticoagulants | 743(1.51%) | 4(0.47%) | 739(1.53%) | 0.11 |
| Antiplatelets | 3217(6.55%) | 9(1.06%) | 3208(6.64%) | 0.29* |
| Mean corpuscular volume, fL | 87.32(7.63);135.4;n=23679 | 87.4(8.6);115.0;n=545 | 87.3(7.6);135.4;n=23134 | 0.01 |
| Basophil, x10^9/L | 0.04(0.07);7.22;n=17089 | 0.0(0.0);0.2;n=408 | 0.0(0.1);7.2;n=16681 | 0.01 |
| Eosinophil, x10^9/L | 0.23(0.27);16.3;n=19223 | 0.2(0.2);2.3;n=442 | 0.2(0.3);16.3;n=18781 | 0.04 |
| Lymphocyte, x10^9/L | 1.98(1.0);45.35;n=19245 | 1.8(0.7);7.5;n=442 | 2.0(1.0);45.4;n=18803 | 0.25* |
| Monocyte, x10^9/L | 0.53(0.25);5.93;n=19245 | 0.6(0.3);2.6;n=442 | 0.5(0.2);5.9;n=18803 | 0.14 |
| Neutrophil, x10^9/L | 5.28(2.77);72.2;n=19245 | 5.5(2.8);23.4;n=442 | 5.3(2.8);72.2;n=18803 | 0.06 |
| White blood cells, x10^9/L | 7.96(3.06);180.4;n=23686 | 8.1(2.9);25.5;n=545 | 8.0(3.1);180.4;n=23141 | 0.03 |
| Mean cell haemoglobin, pg | 29.32(3.01);46.1;n=23679 | 29.2(3.4);39.5;n=545 | 29.3(3.0);46.1;n=23134 | 0.04 |
| Myelocyte, x10^9/L | 0.72(3.37);36.1;n=116 | 0.5(0.6);1.2;n=3 | 0.7(3.4);36.1;n=113 | 0.08 |
| Platelet, x10^9/L | 239.32(72.87);999.0;n=23687 | 217.9(67.7);555.0;n=545 | 239.8(72.9);999.0;n=23142 | 0.31* |
| Reticulocyte, x10^9/L | 65.3(82.27);1735.06;n=496 | 62.2(27.0);123.0;n=20 | 65.4(83.8);1735.1;n=476 | 0.05 |
| Red blood cells, x10^12/L | 4.45(0.7);9.18;n=23679 | 4.3(0.7);7.1;n=545 | 4.5(0.7);9.2;n=23134 | 0.24* |
| Hematocrit, L/L | 0.39(0.05);0.61;n=19699 | 0.41(0.1);0.5;n=449 | 0.38(0.1);0.6;n=19250 | 0.30* |
| Potassium, mmol/L | 4.34(0.49);8.53;n=39895 | 4.4(0.5);6.4;n=763 | 4.3(0.5);8.5;n=39132 | 0.07 |
| Urate, mmol/L | 0.4(0.12);1.27;n=5892 | 0.37(0.1);1.0;n=152 | 0.42(0.1);1.3;n=5740 | 0.33* |
| Albumin, g/L | 41.5(4.14);55.1;n=29670 | 39.9(4.1);51.0;n=617 | 41.5(4.1);55.1;n=29053 | 0.39* |
| Sodium, mmol/L | 139.32(2.97);170.0;n=39913 | 139.3(3.2);149.0;n=763 | 139.3(3.0);170.0;n=39150 | 0.01 |
| Urea, mmol/L | 6.83(3.84);61.12;n=39901 | 8.5(4.6);48.5;n=762 | 6.8(3.8);61.1;n=39139 | 0.40* |
| Protein, g/L | 73.77(5.64);113.0;n=27856 | 73.1(6.4);94.0;n=572 | 73.8(5.6);113.0;n=27284 | 0.12 |
| Creatinine, umol/L | 100.83(88.5);1776.0;n=40019 | 126.7(103.8);1276.0;n=763 | 100.3(88.1);1776.0;n=39256 | 0.27* |
| Alkaline phosphatase, U/L | 77.63(33.18);1679.0;n=29781 | 82.6(45.0);652.0;n=620 | 77.5(32.9);1679.0;n=29161 | 0.13 |
| Aspartate transaminase, U/L | 28.1(60.62);3699.9;n=7950 | 29.4(58.2);538.0;n=165 | 28.1(60.7);3699.9;n=7785 | 0.02 |
| Alanine transaminase, U/L | 27.98(25.01);738.0;n=23744 | 22.4(21.3);350.0;n=491 | 28.1(25.1);738.0;n=23253 | 0.24* |
| Bilirubin, umol/L | 11.08(6.86);439.1;n=29608 | 12.0(6.6);95.0;n=615 | 11.1(6.9);439.1;n=28993 | 0.14 |
| Triglyceride, mmol/L | 1.71(1.41);44.88;n=37464 | 1.6(1.0);6.6;n=688 | 1.7(1.4);44.9;n=36776 | 0.12 |
| Total cholesterol, mmol/L | 4.02(1.32);20.29;n=37493 | 3.8(1.2);10.2;n=689 | 4.0(1.3);20.3;n=36804 | 0.14 |
| LDL, mmol/L | 2.38(0.8);17.76;n=33304 | 2.2(0.8);7.1;n=630 | 2.4(0.8);17.8;n=32674 | 0.24* |
| HDL, mmol/L | 1.19(0.33);5.92;n=33867 | 1.2(0.3);3.5;n=635 | 1.2(0.3);5.9;n=33232 | 0.08 |
| Fasting blood glucose, mmol/L | 8.7(3.72);67.3;n=34303 | 8.8(4.1);34.5;n=643 | 8.7(3.7);67.3;n=33660 | 0.02 |
| HbA1c, g/dL | 10.87(4.98);19.9;n=24139 | 10.4(4.8);18.6;n=559 | 10.9(5.0);19.9;n=23580 | 0.11 |

HF: heart failure; IHD: ischemic heart disease; COPD: chronic obstructive pulmonary disease; LDL: low-density lipoprotein cholesterol; HDL: high-density lipoprotein cholesterol.

**Supplementary Table 3. Baseline characteristics of overall patients and patients with/without new onset stroke.**

* for SMD$\geq$0.2; # indicates the difference between patients with/without new onset stroke.

| **Characteristics** | **Overall (N=49563)**  **Mean(SD);Max;N or Count(%)** | **New Stroke (N=1216) Mean(SD);Max;N or Count(%)** | **No New Stroke (N=48347) Mean(SD);Max;N or Count(%)** | **SMD^#^** |
| --- | --- | --- | --- | --- |
| Male gender | 27244(54.96%) | 629(51.72%) | 26615(55.04%) | 0.07 |
| Baseline age, year | 66.74(12.97);104.55;n=49563 | 74.2(10.6);102.3;n=1216 | 66.6(13.0);104.5;n=48347 | 0.65* |
| <50 | 4595(9.27%) | 22(1.80%) | 4573(9.45%) | 0.34* |
| [50-60] | 10432(21.04%) | 100(8.22%) | 10332(21.37%) | 0.38* |
| [60-70] | 14998(30.26%) | 289(23.76%) | 14709(30.42%) | 0.15 |
| [70-80] | 10870(21.93%) | 402(33.05%) | 10468(21.65%) | 0.26* |
| >80 | 8668(17.48%) | 403(33.14%) | 8265(17.09%) | 0.38* |
| Cardiovascular | 13095(26.42%) | 460(37.82%) | 12635(26.13%) | 0.25* |
| Respiratory | 12715(25.65%) | 462(37.99%) | 12253(25.34%) | 0.27* |
| Renal | 11861(23.93%) | 455(37.41%) | 11406(23.59%) | 0.30* |
| Endocrine | 1055(2.12%) | 38(3.12%) | 1017(2.10%) | 0.06 |
| Hypertension | 26376(53.21%) | 1034(85.03%) | 25342(52.41%) | 0.75* |
| Gastrointestinal | 13540(27.31%) | 472(38.81%) | 13068(27.02%) | 0.25* |
| HF | 854(1.72%) | 20(1.64%) | 834(1.72%) | 0.01 |
| IHD | 5296(10.68%) | 118(9.70%) | 5178(10.71%) | 0.03 |
| Dementia | 569(1.14%) | 22(1.80%) | 547(1.13%) | 0.06 |
| COPD | 11(0.02%) | 0(0.00%) | 11(0.02%) | 0.02 |
| Cancer | 1457(2.93%) | 35(2.87%) | 1422(2.94%) | <0.01 |
| Charlson score | 2.57(1.75);15.0;n=49563 | 3.4(1.6);14.0;n=1216 | 2.5(1.8);15.0;n=48347 | 0.48* |
| SGLT2I | 12482(25.18%) | 39(3.20%) | 12443(25.73%) | 0.68* |
| DPP4I | 37081(74.81%) | 1177(96.79%) | 35904(74.26%) | 0.68* |
| Gliclazide | 33581(67.75%) | 958(78.78%) | 32623(67.47%) | 0.26* |
| Glimepiride | 6355(12.82%) | 152(12.50%) | 6203(12.83%) | 0.01 |
| Metformin | 34430(69.46%) | 902(74.17%) | 33528(69.34%) | 0.11 |
| Sulphonylurea | 4898(9.88%) | 116(9.53%) | 4782(9.89%) | 0.01 |
| Insulin | 10960(22.11%) | 259(21.29%) | 10701(22.13%) | 0.02 |
| Thiozolidinedone | 857(1.72%) | 10(0.82%) | 847(1.75%) | 0.08 |
| Meglitinide | 4407(8.89%) | 105(8.63%) | 4302(8.89%) | 0.01 |
| Glucagon-like peptide-1 agonist | 195(0.39%) | 1(0.08%) | 194(0.40%) | 0.06 |
| Acarbose | 384(0.77%) | 8(0.65%) | 376(0.77%) | 0.01 |
| Diuretics for hypertension | 630(1.27%) | 3(0.24%) | 627(1.29%) | 0.12 |
| Diuretics for HF | 740(1.49%) | 1(0.08%) | 739(1.52%) | 0.16 |
| Anticoagulants | 830(1.67%) | 5(0.41%) | 825(1.70%) | 0.13 |
| Antiplatelets | 3098(6.25%) | 18(1.48%) | 3080(6.37%) | 0.25* |
| Lipid-lowering drugs | 7143(14.41%) | 31(2.54%) | 7112(14.71%) | 0.44* |
| Mean corpuscular volume, fL | 87.37(7.64);135.4;n=24214 | 87.5(7.7);119.9;n=621 | 87.4(7.6);135.4;n=23593 | 0.02 |
| Basophil, x10^9/L | 0.04(0.07);7.22;n=17508 | 0.0(0.0);0.2;n=419 | 0.0(0.1);7.2;n=17089 | 0.04 |
| Eosinophil, x10^9/L | 0.23(0.27);16.3;n=19725 | 0.2(0.2);2.3;n=487 | 0.2(0.3);16.3;n=19238 | 0.01 |
| Lymphocyte, x10^9/L | 1.97(1.0);45.35;n=19747 | 1.8(0.9);8.3;n=487 | 2.0(1.0);45.4;n=19260 | 0.16 |
| Monocyte, x10^9/L | 0.53(0.25);5.93;n=19747 | 0.5(0.2);1.9;n=487 | 0.5(0.2);5.9;n=19260 | 0.09 |
| Neutrophil, x10^9/L | 5.29(2.79);72.2;n=19747 | 5.6(2.9);21.8;n=487 | 5.3(2.8);72.2;n=19260 | 0.13 |
| White blood cells, x10^9/L | 7.95(3.08);180.4;n=24222 | 8.3(4.5);94.6;n=621 | 7.9(3.0);180.4;n=23601 | 0.08 |
| Mean cell haemoglobin, pg | 29.32(3.01);46.1;n=24214 | 29.3(3.0);41.0;n=621 | 29.3(3.0);46.1;n=23593 | 0.01 |
| Myelocyte, x10^9/L | 0.66(3.2);36.1;n=129 | 0.3(0.2);0.5;n=4 | 0.7(3.2);36.1;n=125 | 0.14 |
| Platelet, x10^9/L | 237.79(72.88);999.0;n=24223 | 234.7(72.8);582.0;n=621 | 237.9(72.9);999.0;n=23602 | 0.04 |
| Reticulocyte, x10^9/L | 65.31(80.08);1735.06;n=527 | 65.0(30.4);143.0;n=15 | 65.3(81.1);1735.1;n=512 | 0.01 |
| Red blood cells, x10^12/L | 4.44(0.71);9.18;n=24214 | 4.3(0.7);6.6;n=621 | 4.4(0.7);9.2;n=23593 | 0.25* |
| Hematocrit, L/L | 0.38(0.05);0.61;n=20177 | 0.41(0.1);0.5;n=527 | 0.4(0.1);0.6;n=19650 | 0.28* |
| Potassium, mmol/L | 4.34(0.49);8.53;n=40439 | 4.4(0.6);8.1;n=1071 | 4.3(0.5);8.5;n=39368 | 0.11 |
| Urate, mmol/L | 0.41(0.12);1.27;n=6080 | 0.37(0.1);0.9;n=150 | 0.42(0.1);1.3;n=5930 | 0.16 |
| Albumin, g/L | 41.43(4.17);55.1;n=30204 | 40.5(4.2);53.1;n=737 | 41.5(4.2);55.1;n=29467 | 0.22* |
| Sodium, mmol/L | 139.32(2.99);170.0;n=40460 | 139.3(3.3);149.0;n=1074 | 139.3(3.0);170.0;n=39386 | 0.01 |
| Urea, mmol/L | 6.89(3.9);61.12;n=40445 | 7.7(4.6);49.5;n=1077 | 6.9(3.9);61.1;n=39368 | 0.19 |
| Protein, g/L | 73.74(5.67);113.0;n=28365 | 73.4(6.0);94.0;n=689 | 73.8(5.7);113.0;n=27676 | 0.06 |
| Creatinine, umol/L | 101.61(89.15);1776.0;n=40567 | 113.6(88.0);967.0;n=1078 | 101.3(89.2);1776.0;n=39489 | 0.14 |
| Alkaline phosphatase, U/L | 77.86(33.77);1679.0;n=30315 | 79.5(32.2);377.2;n=739 | 77.8(33.8);1679.0;n=29576 | 0.05 |
| Aspartate transaminase, U/L | 27.83(54.99);3699.9;n=8124 | 33.1(65.8);730.5;n=193 | 27.7(54.7);3699.9;n=7931 | 0.09 |
| Alanine transaminase, U/L | 27.85(24.96);738.0;n=24217 | 24.3(23.7);287.0;n=621 | 27.9(25.0);738.0;n=23596 | 0.15 |
| Bilirubin, umol/L | 11.16(6.93);439.1;n=30142 | 11.0(7.2);102.5;n=735 | 11.2(6.9);439.1;n=29407 | 0.02 |
| Triglyceride, mmol/L | 1.7(1.39);44.88;n=37877 | 1.6(1.2);13.3;n=990 | 1.7(1.4);44.9;n=36887 | 0.05 |
| Total cholesterol, mmol/L | 4.01(1.31);14.8;n=37909 | 3.9(1.3);10.2;n=990 | 4.0(1.3);14.8;n=36919 | 0.10 |
| LDL, mmol/L | 2.37(0.79);11.98;n=33725 | 2.3(0.8);6.4;n=872 | 2.4(0.8);12.0;n=32853 | 0.11 |
| HDL, mmol/L | 1.19(0.33);5.92;n=34283 | 1.2(0.3);3.7;n=885 | 1.2(0.3);5.9;n=33398 | 0.01 |
| Fasting blood glucose, mmol/L | 8.69(3.76);78.33;n=34781 | 8.6(3.5);26.5;n=917 | 8.7(3.8);78.3;n=33864 | 0.02 |
| HbA1c, g/dL | 10.86(4.97);19.9;n=24672 | 10.9(4.4);18.3;n=652 | 10.9(5.0);19.9;n=24020 | <0.01 |

HF: heart failure; IHD: ischemic heart disease; COPD: chronic obstructive pulmonary disease; LDL: low-density lipoprotein cholesterol; HDL: high-density lipoprotein cholesterol.

**Table 2. Baseline characteristics among overall, all-cause mortality and cardiovascular mortality patients in the HF cohort.**

* for SMD≤ 0.2; # indicates the difference between all-cause mortality and cardiovascular mortality patients

| **Characteristics** | **Overall (N=48875)**  **Mean(SD);Max;N or Count(%)** | **All-cause mortality (N=5993) Mean(SD);Max;N or Count(%)** | **Cardiovascular mortality (N=803)**  **Mean(SD);Max;N or Count(%)** | **SMD^#^** |
| --- | --- | --- | --- | --- |
| New HF | 1142(2.33%) | 915(15.26%) | 210(26.15%) | 0.27 |
| Male gender | 26739(54.70%) | 3231(53.91%) | 424(52.80%) | 0.02* |
| Baseline age, year | 66.97(13.04);104.55;n=48875 | 78.04(11.41);103.97;n=5993 | 76.47(11.88);100.91;n=803 | 0.13* |
| Overall admission times | 157.52(390.81);2740.0;n=43388 | 127.12(337.52);2627.0;n=5073 | 112.49(312.88);2623.0;n=685 | 0.04* |
| Overall hospital stay, day | 7857.53(22118.01);169749.0;n=43388 | 6561.71(19385.21);169749.0;n=5073 | 6263.55(19610.8);169749.0;n=685 | 0.02* |
| Emergency readmission times | 42.26(109.62);1181.0;n=23522 | 32.89(81.33);1181.0;n=2511 | 33.41(94.49);1181.0;n=344 | 0.01* |
| Cardiovascular | 12520(25.61%) | 3610(60.23%) | 694(86.42%) | 0.62 |
| Respiratory | 12528(25.63%) | 4359(72.73%) | 574(71.48%) | 0.03* |
| Renal | 11878(24.30%) | 3508(58.53%) | 500(62.26%) | 0.08* |
| Endocrine | 986(2.01%) | 351(5.85%) | 18(2.24%) | 0.18* |
| Hypertension | 26554(54.33%) | 4216(70.34%) | 543(67.62%) | 0.06* |
| Gastrointestinal | 13547(27.71%) | 3186(53.16%) | 331(41.22%) | 0.24 |
| Stroke | 8719(17.83%) | 2200(36.70%) | 366(45.57%) | 0.18* |
| IHD | 4753(9.72%) | 1165(19.43%) | 283(35.24%) | 0.36 |
| SCD | 1578(3.22%) | 538(8.97%) | 152(18.92%) | 0.29 |
| Dementia and Alzheimer | 599(1.22%) | 254(4.23%) | 13(1.61%) | 0.16* |
| COPD | 10(0.02%) | 4(0.06%) | 0(0.00%) | 0.04* |
| Cancer | 1353(2.76%) | 511(8.52%) | 28(3.48%) | 0.21 |
| Charlson score | 2.6(1.74);15.0;n=48875 | 4.52(2.07);15.0;n=5993 | 4.3(1.84);11.0;n=803 | 0.11* |
| SGLT2 | 11011(22.52%) | 207(3.45%) | 33(4.10%) | 0.03* |
| DPP4i | 37864(77.47%) | 5786(96.54%) | 770(95.89%) | 0.03* |
| Gliclazide | 33392(68.32%) | 4190(69.91%) | 549(68.36%) | 0.03* |
| Glimepiride | 6271(12.83%) | 563(9.39%) | 87(10.83%) | 0.05* |
| Metformin | 33804(69.16%) | 3294(54.96%) | 445(55.41%) | 0.01* |
| Sulphonylurea | 4803(9.82%) | 568(9.47%) | 78(9.71%) | 0.01* |
| Insulin | 10615(21.71%) | 1789(29.85%) | 272(33.87%) | 0.09* |
| Thiozolidinedone | 792(1.62%) | 48(0.80%) | 7(0.87%) | 0.01* |
| Meglitinide | 4310(8.81%) | 513(8.55%) | 66(8.21%) | 0.01* |
| Glucagon-like peptide-1 agonist | 163(0.33%) | 3(0.05%) | 1(0.12%) | 0.03* |
| Acarbose | 378(0.77%) | 74(1.23%) | 9(1.12%) | 0.01* |
| Diuretics for hypertension | 512(1.04%) | 11(0.18%) | 1(0.12%) | 0.02* |
| Lipid-lowering drugs | 5918(12.10%) | 106(1.76%) | 18(2.24%) | 0.03* |
| Mean corpuscular volume, fL | 87.41(7.64);135.4;n=23539 | 88.23(8.12);124.7;n=4017 | 88.14(7.85);112.4;n=557 | 0.01* |
| Basophil, x10^9/L | 0.04(0.07);7.22;n=17058 | 0.03(0.04);0.83;n=3141 | 0.03(0.04);0.2;n=449 | 0.05* |
| Eosinophil, x10^9/L | 0.22(0.26);16.3;n=19144 | 0.23(0.27);5.05;n=3498 | 0.23(0.21);1.72;n=492 | 0.0* |
| Lymphocyte, x10^9/L | 1.95(0.98);45.35;n=19165 | 1.64(0.95);30.24;n=3506 | 1.71(0.78);5.61;n=492 | 0.08* |
| Metamyelocyte, x10^9/L | 0.75(3.26);22.49;n=47 | 0.17(0.14);0.49;n=18 | 0.11(0.01);0.11;n=2 | 0.61 |
| Monocyte, x10^9/L | 0.52(0.25);5.93;n=19165 | 0.56(0.31);5.93;n=3506 | 0.54(0.25);2.4;n=492 | 0.05* |
| Neutrophil, x10^9/L | 5.3(2.81);72.2;n=19165 | 5.87(3.34);32.74;n=3506 | 5.75(2.76);22.74;n=492 | 0.04* |
| White blood count, x10^9/L | 7.95(3.09);180.4;n=23547 | 8.21(3.57);94.6;n=4017 | 8.28(4.56);94.6;n=557 | 0.02* |
| Mean cell haemoglobin, pg | 29.34(3.01);46.1;n=23539 | 29.48(3.18);45.5;n=4017 | 29.41(3.01);37.1;n=557 | 0.02* |
| Myelocyte, x10^9/L | 0.66(3.25);36.1;n=124 | 0.35(0.26);1.06;n=50 | 0.27(0.16);0.46;n=4 | 0.35 |
| Platelet, x10^9/L | 237.93(73.21);999.0;n=23548 | 225.8(81.03);854.0;n=4017 | 227.99(78.49);588.0;n=557 | 0.03* |
| Reticulocyte, x10^9/L | 65.1(80.43);1735.06;n=524 | 69.46(131.28);1735.06;n=172 | 61.43(30.04);150.4;n=28 | 0.08* |
| Red blood count, x10^12/L | 4.43(0.71);9.18;n=23539 | 4.04(0.73);8.04;n=4017 | 4.09(0.76);6.9;n=557 | 0.06* |
| Hematocrit, L/L | 0.38(0.05);0.61;n=19537 | 0.35(0.06);0.59;n=3317 | 0.36(0.06);0.59;n=447 | 0.07* |
| K/Potassium, mmol/L | 4.34(0.49);8.53;n=39605 | 4.36(0.58);8.1;n=5132 | 4.39(0.57);6.84;n=696 | 0.05* |
| Urate, mmol/L | 0.4(0.12);1.27;n=5891 | 0.44(0.13);1.08;n=978 | 0.47(0.14);0.9;n=165 | 0.25 |
| Albumin, g/L | 41.38(4.19);55.1;n=29365 | 38.15(4.98);52.0;n=4300 | 38.5(4.91);50.8;n=592 | 0.07* |
| Na/Sodium, mmol/L | 139.31(3.0);170.0;n=39627 | 138.74(3.86);170.0;n=5137 | 138.68(4.02);170.0;n=697 | 0.01* |
| Urea, mmol/L | 6.92(3.94);61.12;n=39612 | 9.55(5.99);61.12;n=5135 | 10.02(5.91);36.1;n=697 | 0.08* |
| Protein, g/L | 73.71(5.67);113.0;n=27559 | 72.09(6.92);104.0;n=4050 | 72.35(6.72);103.0;n=557 | 0.04* |
| Creatinine, umol/L | 102.33(90.66);1776.0;n=39731 | 155.01(155.79);1584.0;n=5140 | 166.47(178.89);1498.0;n=697 | 0.07* |
| Alkaline phosphatase, U/L | 77.93(33.82);1679.0;n=29474 | 89.86(51.24);1679.0;n=4310 | 88.59(38.28);381.0;n=594 | 0.03* |
| Aspartate transaminase, U/L | 28.14(61.86);3699.9;n=7762 | 30.8(55.15);892.0;n=1189 | 26.78(42.11);523.2;n=169 | 0.08* |
| Alanine transaminase, U/L | 27.58(24.97);738.0;n=23450 | 22.46(25.23);486.0;n=3342 | 24.07(28.21);350.0;n=460 | 0.06* |
| Bilirubin, umol/L | 11.11(6.57);439.1;n=29302 | 10.76(8.14);221.8;n=4295 | 10.42(6.01);75.0;n=592 | 0.05* |
| Triglyceride, mmol/L | 1.7(1.4);44.88;n=37061 | 1.62(1.18);19.13;n=4319 | 1.67(1.22);15.9;n=606 | 0.04* |
| Total cholesterol, mmol/L | 4.01(1.32);20.29;n=37090 | 3.98(1.34);20.29;n=4328 | 4.06(1.3);9.3;n=611 | 0.06* |
| Low-density lipoprotein (LDL), mmol/L | 2.37(0.8);17.76;n=32918 | 2.32(0.88);17.76;n=3897 | 2.33(0.89);6.95;n=560 | 0.01* |
| High-density lipoprotein (LDL), mmol/L | 1.2(0.33);5.92;n=33458 | 1.19(0.37);4.4;n=3958 | 1.15(0.35);2.92;n=568 | 0.1* |
| Glucose | 8.69(3.74);78.33;n=34118 | 9.11(4.87);78.33;n=4401 | 9.1(4.49);33.49;n=613 | 0.0* |
| HbA1C, g/dL | 10.8(4.98);19.8;n=24009 | 9.88(4.64);19.1;n=4060 | 9.71(4.94);19.1;n=562 | 0.04* |

**Table 3. Characteristics among overall, all-cause mortality and cardiovascular mortality patients after propensity score matching (1:2) in the HF cohort.**

* for SMD≤ 0.2; # indicates the difference between all-cause mortality and cardiovascular mortality patients

| **Characteristics** | **Overall (N=33033)**  **Mean(SD);Max;N or Count(%)** | **All-cause mortality (N=1231) Mean(SD);Max;N or Count(%)** | **Cardiovascular mortality (N=213) Mean(SD);Max;N or Count(%)** | **SMD^#^** |
| --- | --- | --- | --- | --- |
| New HF | 246(0.74%) | 184(14.94%) | 46(21.59%) | 0.17* |
| Male gender | 20451(61.91%) | 812(65.96%) | 137(64.31%) | 0.03* |
| Baseline age, year | 60.25(11.32);96.55;n=33033 | 68.52(11.71);96.55;n=1231 | 68.53(11.75);96.24;n=213 | 0.0* |
| Overall admission times | 159.73(391.14);2740.0;n=29530 | 130.53(348.79);2361.0;n=1038 | 89.81(178.48);1219.0;n=183 | 0.15* |
| Overall hospital stay, day | 7891.77(22032.25);169749.0;n=29530 | 7278.28(21452.15);153095.0;n=1038 | 5053.87(13130.49);70805.0;n=183 | 0.13* |
| Emergency readmission times | 43.03(112.9);1181.0;n=16333 | 31.12(77.72);709.0;n=507 | 25.44(47.99);227.0;n=104 | 0.09* |
| Cardiovascular | 5578(16.88%) | 567(46.06%) | 158(74.17%) | 0.6 |
| Respiratory | 5667(17.15%) | 685(55.64%) | 119(55.86%) | 0.00* |
| Renal | 2370(7.17%) | 248(20.14%) | 42(19.71%) | 0.01* |
| Endocrine | 441(1.33%) | 69(5.60%) | 3(1.40%) | 0.23 |
| Hypertension | 9726(29.44%) | 536(43.54%) | 91(42.72%) | 0.02* |
| Gastrointestinal | 5053(15.29%) | 425(34.52%) | 36(16.90%) | 0.41 |
| Stroke | 4076(12.33%) | 330(26.80%) | 82(38.49%) | 0.25 |
| IHD | 3271(9.90%) | 233(18.92%) | 73(34.27%) | 0.35 |
| SCD | 848(2.56%) | 84(6.82%) | 29(13.61%) | 0.23 |
| Dementia and Alzheimer | 21(0.06%) | 4(0.32%) | 0(0.00%) | 0.08* |
| COPD | 6(0.01%) | 2(0.16%) | 0(0.00%) | 0.06* |
| Cancer | 514(1.55%) | 85(6.90%) | 7(3.28%) | 0.17* |
| Charlson score | 1.72(1.21);12.0;n=33033 | 2.89(1.72);12.0;n=1231 | 2.94(1.51);8.0;n=213 | 0.03* |
| SGLT2 | 11011(33.33%) | 207(16.81%) | 33(15.49%) | 0.04* |
| DPP4i | 22022(66.66%) | 1024(83.18%) | 180(84.50%) | 0.04* |
| Gliclazide | 17935(54.29%) | 741(60.19%) | 143(67.13%) | 0.14* |
| Glimepiride | 4122(12.47%) | 138(11.21%) | 20(9.38%) | 0.06* |
| Metformin | 20934(63.37%) | 811(65.88%) | 140(65.72%) | 0.00* |
| Sulphonylurea | 3092(9.36%) | 103(8.36%) | 14(6.57%) | 0.07* |
| Insulin | 9408(28.48%) | 416(33.79%) | 79(37.08%) | 0.07* |
| Thiozolidinedone | 1434(4.34%) | 43(3.49%) | 6(2.81%) | 0.04* |
| Meglitinide | 2892(8.75%) | 100(8.12%) | 13(6.10%) | 0.08* |
| Glucagon-like peptide-1 agonist | 438(1.32%) | 17(1.38%) | 1(0.46%) | 0.10* |
| Acarbose | 234(0.70%) | 16(1.29%) | 3(1.40%) | 0.01* |
| Diuretics for hypertension | 512(1.54%) | 11(0.89%) | 1(0.46%) | 0.05* |
| Lipid-lowering drugs | 5948(18.00%) | 107(8.69%) | 19(8.92%) | 0.01* |
| Mean corpuscular volume, fL | 86.76(7.47);134.3;n=14900 | 87.65(8.1);112.9;n=710 | 87.02(7.53);106.9;n=129 | 0.08* |
| Basophil, x10^9/L | 0.04(0.08);7.22;n=10451 | 0.04(0.04);0.2;n=550 | 0.05(0.05);0.2;n=97 | 0.18* |
| Eosinophil, x10^9/L | 0.22(0.21);4.11;n=11941 | 0.22(0.21);1.15;n=607 | 0.23(0.22);1.1;n=112 | 0.07* |
| Lymphocyte, x10^9/L | 2.12(0.93);34.8;n=11945 | 1.77(0.88);6.2;n=608 | 1.92(0.93);5.0;n=112 | 0.16* |
| Metamyelocyte, x10^9/L | 1.87(5.53);22.49;n=16 | 0.05(nan);0.05;n=1 | - | - |
| Monocyte, x10^9/L | 0.51(0.22);3.6;n=11945 | 0.54(0.25);2.4;n=608 | 0.55(0.3);2.4;n=112 | 0.05* |
| Neutrophil, x10^9/L | 5.03(2.46);72.2;n=11945 | 5.66(2.99);23.5;n=608 | 5.8(2.59);19.8;n=112 | 0.05* |
| White blood count, x10^9/L | 7.88(3.27);180.4;n=14908 | 8.62(7.07);94.6;n=710 | 10.92(15.23);94.6;n=129 | 0.19* |
| Mean cell haemoglobin, pg | 29.19(2.96);43.8;n=14900 | 29.32(3.21);36.8;n=710 | 29.06(2.96);36.1;n=129 | 0.08* |
| Myelocyte, x10^9/L | 0.93(4.67);36.1;n=59 | 0.28(0.15);0.67;n=18 | 0.46(nan);0.46;n=1 | nan |
| Platelet, x10^9/L | 244.68(70.9);783.0;n=14905 | 234.65(83.27);667.0;n=710 | 247.91(101.82);588.0;n=129 | 0.14* |
| Reticulocyte, x10^9/L | 69.01(29.27);175.0;n=197 | 67.22(25.79);103.4;n=16 | 64.96(25.95);99.8;n=6 | 0.09* |
| Red blood count, x10^12/L | 4.67(0.63);9.18;n=14900 | 4.43(0.75);7.23;n=710 | 4.44(0.81);6.9;n=129 | 0.02* |
| Hematocrit, L/L | 0.4(0.05);0.61;n=12220 | 0.38(0.06);0.57;n=599 | 0.38(0.07);0.57;n=102 | 0.04* |
| K/Potassium, mmol/L | 4.31(0.44);7.61;n=24234 | 4.34(0.51);7.61;n=1003 | 4.33(0.49);6.0;n=169 | 0.03* |
| Urate, mmol/L | 0.39(0.11);0.97;n=3832 | 0.42(0.14);0.97;n=133 | 0.48(0.12);0.74;n=34 | 0.46 |
| Albumin, g/L | 42.42(3.55);55.1;n=19125 | 39.9(4.85);50.1;n=800 | 39.88(4.68);48.6;n=145 | 0.01* |
| Na/Sodium, mmol/L | 139.41(2.8);170.0;n=24240 | 138.9(3.66);170.0;n=1004 | 138.37(4.5);170.0;n=169 | 0.13* |
| Urea, mmol/L | 6.06(2.64);44.6;n=24234 | 7.17(3.75);36.1;n=1004 | 7.35(4.66);36.1;n=169 | 0.04* |
| Protein, g/L | 74.19(5.21);113.0;n=17915 | 72.89(7.31);103.0;n=750 | 74.15(6.58);103.0;n=137 | 0.18* |
| Creatinine, umol/L | 85.9(49.16);1294.0;n=24296 | 106.01(83.59);1276.0;n=1004 | 117.58(127.61);1276.0;n=169 | 0.11* |
| Alkaline phosphatase, U/L | 75.15(28.37);744.0;n=19184 | 86.7(37.4);446.0;n=806 | 88.18(32.15);286.0;n=147 | 0.04* |
| Aspartate transaminase, U/L | 26.52(26.41);701.0;n=4627 | 33.38(45.27);292.0;n=206 | 28.72(30.49);171.4;n=41 | 0.12* |
| Alanine transaminase, U/L | 30.43(27.31);738.0;n=15144 | 30.68(40.38);303.0;n=638 | 32.04(35.04);182.0;n=112 | 0.04* |
| Bilirubin, umol/L | 11.17(5.62);97.0;n=19093 | 11.63(9.42);93.5;n=799 | 12.72(11.54);75.0;n=145 | 0.1* |
| Triglyceride, mmol/L | 1.76(1.52);31.0;n=23010 | 1.62(1.05);12.7;n=893 | 1.6(1.18);12.7;n=150 | 0.02* |
| Total cholesterol, mmol/L | 4.18(1.27);14.8;n=23023 | 4.1(1.39);8.9;n=894 | 4.21(1.11);7.03;n=151 | 0.09* |
| Low-density lipoprotein (LDL), mmol/L | 2.44(0.82);11.98;n=20998 | 2.45(0.87);6.29;n=811 | 2.4(0.77);4.54;n=146 | 0.06* |
| High-density lipoprotein (LDL), mmol/L | 1.19(0.33);5.92;n=21440 | 1.18(0.34);2.61;n=819 | 1.13(0.34);2.31;n=146 | 0.14* |
| Glucose | 8.77(3.87);64.6;n=21229 | 9.33(4.78);39.5;n=870 | 9.71(5.78);33.49;n=154 | 0.07* |
| HbA1C, g/dL | 11.19(5.26);19.8;n=15140 | 11.05(4.74);19.1;n=718 | 10.16(5.47);19.1;n=129 | 0.17* |

**Table 4. Baseline characteristics among overall, all-cause mortality and cardiovascular mortality patients in the MI cohort.**

* for SMD≤ 0.2; # indicates the difference between all-cause mortality and cardiovascular mortality patients

| **Characteristics** | **All (N=49508)**  **Mean(SD);Max;N or Count(%)** | **Mortality (N=5789) Mean(SD);Max;N or Count(%)** | **Cardiovascular mortality (N=702) Mean(SD);Max;N or Count(%)** | **SMD^#^** |
| --- | --- | --- | --- | --- |
| New MI | 890(1.79%) | 551(9.51%) | 259(36.89%) | 0.69 |
| Male gender | 27136(54.81%) | 3146(54.34%) | 381(54.27%) | 0.00* |
| Baseline age, year | 66.74(12.95);104.55;n=49508 | 77.76(11.44);103.97;n=5789 | 75.92(12.08);100.91;n=702 | 0.16* |
| Overall admission times | 158.95(392.95);2740.0;n=44010 | 123.89(329.6);2627.0;n=4902 | 99.15(279.51);2591.0;n=599 | 0.08* |
| Overall hospital stay, day | 7922.51(22189.54);169749.0;n=44009 | 6356.47(18858.53);169749.0;n=4901 | 5325.52(17162.92);169749.0;n=599 | 0.06* |
| Emergency readmission times | 42.54(109.7);1181.0;n=23941 | 32.42(77.81);931.0;n=2412 | 28.94(73.38);931.0;n=299 | 0.05* |
| Cardiovascular | 12211(24.66%) | 3332(57.55%) | 587(83.61%) | 0.6 |
| Respiratory | 12426(25.09%) | 4187(72.32%) | 495(70.51%) | 0.04* |
| Renal | 11605(23.44%) | 3310(57.17%) | 427(60.82%) | 0.07* |
| Endocrine | 1035(2.09%) | 369(6.37%) | 13(1.85%) | 0.23 |
| Hypertension | 26543(53.61%) | 4016(69.37%) | 463(65.95%) | 0.07* |
| Gastrointestinal | 13475(27.21%) | 3060(52.85%) | 279(39.74%) | 0.27 |
| Stroke | 8549(17.26%) | 1990(34.37%) | 291(41.45%) | 0.15* |
| HF | 719(1.45%) | 307(5.30%) | 50(7.12%) | 0.08* |
| IHD | 3768(7.61%) | 724(12.50%) | 157(22.36%) | 0.26 |
| SCD | 42(0.08%) | 21(0.36%) | 6(0.85%) | 0.06* |
| Dementia and Alzheimer | 585(1.18%) | 245(4.23%) | 12(1.70%) | 0.15* |
| COPD | 12(0.02%) | 5(0.08%) | 0(0.00%) | 0.04* |
| Cancer | 1469(2.96%) | 590(10.19%) | 25(3.56%) | 0.26 |
| Charlson score | 2.55(1.73);18.0;n=49508 | 4.49(2.18);18.0;n=5789 | 3.98(1.74);14.0;n=702 | 0.26 |
| SGLT2 | 12391(25.02%) | 249(4.30%) | 49(6.98%) | 0.12* |
| DPP4i | 37117(74.97%) | 5540(95.69%) | 653(93.01%) | 0.12* |
| Gliclazide | 33624(67.91%) | 4072(70.34%) | 486(69.23%) | 0.02* |
| Glimepiride | 6350(12.82%) | 550(9.50%) | 74(10.54%) | 0.03* |
| Metformin | 34544(69.77%) | 3256(56.24%) | 411(58.54%) | 0.05* |
| Sulphonylurea | 4887(9.87%) | 555(9.58%) | 65(9.25%) | 0.01* |
| Insulin | 10836(21.88%) | 1673(28.89%) | 230(32.76%) | 0.08* |
| Thiozolidinedone | 864(1.74%) | 51(0.88%) | 8(1.13%) | 0.03* |
| Meglitinide | 4395(8.87%) | 508(8.77%) | 57(8.11%) | 0.02* |
| Glucagon-like peptide-1 agonist | 196(0.39%) | 3(0.05%) | 1(0.14%) | 0.03* |
| Acarbose | 397(0.80%) | 71(1.22%) | 9(1.28%) | 0.00* |
| Diuretics for hypertension | 641(1.29%) | 14(0.24%) | 1(0.14%) | 0.02* |
| Diuretics for heart failure | 710(1.43%) | 40(0.69%) | 16(2.27%) | 0.13* |
| Lipid-lowering drugs | 7073(14.28%) | 141(2.43%) | 29(4.13%) | 0.10* |
| Mean corpuscular volume, fL | 87.37(7.64);134.3;n=24129 | 88.22(8.12);124.7;n=3879 | 88.27(7.93);112.4;n=488 | 0.01* |
| Basophil, x10^9/L | 0.04(0.07);7.22;n=17417 | 0.03(0.04);0.83;n=3030 | 0.04(0.04);0.2;n=390 | 0.06* |
| Eosinophil, x10^9/L | 0.22(0.27);16.3;n=19631 | 0.23(0.26);3.2;n=3370 | 0.24(0.24);2.56;n=431 | 0.05* |
| Lymphocyte, x10^9/L | 1.97(0.99);45.35;n=19651 | 1.65(0.96);30.24;n=3377 | 1.73(0.8);5.61;n=431 | 0.09* |
| Metamyelocyte, x10^9/L | 0.74(3.22);22.49;n=48 | 0.16(0.14);0.49;n=17 | 0.11(0.01);0.11;n=2 | 0.55 |
| Monocyte, x10^9/L | 0.52(0.25);5.93;n=19651 | 0.56(0.31);5.93;n=3377 | 0.56(0.25);2.4;n=431 | 0.0* |
| Neutrophil, x10^9/L | 5.27(2.77);72.2;n=19651 | 5.83(3.32);32.74;n=3377 | 5.76(2.85);22.74;n=431 | 0.02* |
| White blood count, x10^9/L | 7.93(3.06);180.4;n=24136 | 8.18(3.56);94.6;n=3879 | 8.32(4.78);94.6;n=488 | 0.03* |
| Mean cell haemoglobin, pg | 29.32(3.01);45.5;n=24129 | 29.47(3.18);45.5;n=3879 | 29.44(3.01);37.1;n=488 | 0.01* |
| Myelocyte, x10^9/L | 0.66(3.29);36.1;n=121 | 0.32(0.26);1.06;n=48 | 0.3(0.15);0.46;n=5 | 0.1* |
| Platelet, x10^9/L | 237.91(72.98);999.0;n=24137 | 225.89(81.62);854.0;n=3879 | 226.29(80.37);588.0;n=488 | 0.0* |
| Reticulocyte, x10^9/L | 66.28(81.93);1735.06;n=501 | 72.39(137.8);1735.06;n=155 | 65.13(28.03);150.4;n=24 | 0.07* |
| Red blood count, x10^12/L | 4.45(0.7);9.18;n=24129 | 4.07(0.73);8.04;n=3879 | 4.14(0.76);6.9;n=488 | 0.09* |
| Hematocrit, L/L | 0.39(0.05);0.61;n=20076 | 0.36(0.06);0.59;n=3214 | 0.36(0.06);0.59;n=391 | 0.12* |
| K/Potassium, mmol/L | 4.34(0.49);8.53;n=40388 | 4.36(0.57);8.1;n=4960 | 4.39(0.56);6.1;n=613 | 0.05* |
| Urate, mmol/L | 0.4(0.12);1.03;n=6041 | 0.44(0.13);1.03;n=919 | 0.47(0.13);0.81;n=138 | 0.26 |
| Albumin, g/L | 41.47(4.13);55.1;n=30124 | 38.31(4.98);52.0;n=4150 | 38.82(4.79);50.8;n=523 | 0.11* |
| Na/Sodium, mmol/L | 139.34(2.98);170.0;n=40408 | 138.75(3.89);170.0;n=4963 | 138.84(3.97);170.0;n=614 | 0.02* |
| Urea, mmol/L | 6.83(3.8);61.12;n=40394 | 9.32(5.78);61.12;n=4961 | 9.5(5.58);36.1;n=614 | 0.03* |
| Protein, g/L | 73.79(5.64);113.0;n=28302 | 72.16(6.93);104.0;n=3913 | 72.67(6.66);103.0;n=488 | 0.08* |
| Creatinine, umol/L | 100.24(85.32);1776.0;n=40515 | 148.37(146.32);1598.3;n=4966 | 153.46(156.95);1498.0;n=614 | 0.03* |
| Alkaline phosphatase, U/L | 77.62(33.17);1679.0;n=30234 | 89.38(50.35);1679.0;n=4159 | 88.16(38.06);381.0;n=525 | 0.03* |
| Aspartate transaminase, U/L | 27.67(44.77);2330.0;n=8098 | 31.47(57.98);892.0;n=1158 | 27.97(45.13);523.2;n=149 | 0.07* |
| Alanine transaminase, U/L | 27.89(25.03);738.0;n=24114 | 22.62(24.89);486.0;n=3235 | 24.58(28.5);350.0;n=405 | 0.07* |
| Bilirubin, umol/L | 11.17(6.74);439.1;n=30061 | 10.88(7.64);152.0;n=4145 | 10.83(6.41);75.0;n=523 | 0.01* |
| Triglyceride, mmol/L | 1.7(1.39);44.88;n=37856 | 1.6(1.14);19.13;n=4161 | 1.65(1.1);12.7;n=542 | 0.04* |
| Total cholesterol, mmol/L | 4.01(1.31);20.29;n=37884 | 3.98(1.33);20.29;n=4168 | 4.07(1.26);9.3;n=545 | 0.08* |
| Low-density lipoprotein (LDL), mmol/L | 2.37(0.8);17.76;n=33724 | 2.31(0.88);17.76;n=3757 | 2.32(0.86);6.95;n=504 | 0.01* |
| High-density lipoprotein (LDL), mmol/L | 1.2(0.33);5.92;n=34282 | 1.19(0.37);4.4;n=3813 | 1.15(0.35);2.92;n=511 | 0.1* |
| Glucose | 8.68(3.71);78.33;n=34722 | 9.1(4.95);78.33;n=4240 | 9.1(4.65);43.1;n=541 | 0.01* |
| HbA1C, g/dL | 10.87(4.97);19.9;n=24598 | 9.96(4.65);19.1;n=3920 | 9.78(5.04);19.1;n=492 | 0.04* |

**Table 5. Characteristics among overall, all-cause mortality and cardiovascular mortality patients after propensity score matching (1:2) in the MI cohort.**

* for SMD≤ 0.2; # indicates the difference between all-cause mortality and cardiovascular mortality patients

| **Characteristics** | **All (N=37173)**  **Mean(SD);Max;N or Count(%)** | **All-cause mortality (N=1522) Mean(SD);Max;N or Count(%)** | **Cardiovascular mortality (N=257) Mean(SD);Max;N or Count(%)** | **SMD** |
| --- | --- | --- | --- | --- |
| New MI | 238(0.64%) | 102(6.70%) | 64(24.90%) | 0.52 |
| Male gender | 22929(61.68%) | 1011(66.42%) | 166(64.59%) | 0.04* |
| Baseline age, year | 60.76(11.3);96.24;n=37173 | 68.83(11.58);96.24;n=1522 | 67.83(11.69);96.24;n=257 | 0.09* |
| Overall admission times | 160.25(391.78);2740.0;n=33225 | 131.47(346.16);2361.0;n=1281 | 82.54(165.18);1292.0;n=222 | 0.18* |
| Overall hospital stay, day | 7932.75(22043.5);169749.0;n=33225 | 7384.46(21249.73);153095.0;n=1281 | 4973.11(13796.5);80693.0;n=222 | 0.13* |
| Emergency readmission times | 42.31(110.67);1181.0;n=18381 | 31.93(74.56);709.0;n=632 | 22.49(38.81);203.0;n=126 | 0.16* |
| Cardiovascular | 7016(18.87%) | 763(50.13%) | 196(76.26%) | 0.56 |
| Respiratory | 6624(17.81%) | 880(57.81%) | 146(56.80%) | 0.02* |
| Renal | 2852(7.67%) | 328(21.55%) | 59(22.95%) | 0.03* |
| Endocrine | 543(1.46%) | 85(5.58%) | 2(0.77%) | 0.28 |
| Hypertension | 11436(30.76%) | 689(45.26%) | 110(42.80%) | 0.05* |
| Gastrointestinal | 5827(15.67%) | 498(32.72%) | 41(15.95%) | 0.4 |
| Stroke | 5223(14.05%) | 454(29.82%) | 100(38.91%) | 0.19* |
| HF | 415(1.11%) | 91(5.97%) | 16(6.22%) | 0.01* |
| IHD | 3636(9.78%) | 277(18.19%) | 79(30.73%) | 0.29 |
| SCD | 18(0.04%) | 8(0.52%) | 4(1.55%) | 0.10* |
| Dementia and Alzheimer | 27(0.07%) | 6(0.39%) | 1(0.38%) | 0.00* |
| COPD | 6(0.01%) | 2(0.13%) | 0(0.00%) | 0.05* |
| Cancer | 616(1.65%) | 106(6.96%) | 6(2.33%) | 0.22 |
| Charlson score | 1.75(1.21);12.0;n=37173 | 2.93(1.75);12.0;n=1522 | 2.74(1.34);7.0;n=257 | 0.12* |
| SGLT2 | 12391(33.33%) | 249(16.36%) | 49(19.06%) | 0.07* |
| DPP4i | 24782(66.66%) | 1273(83.63%) | 208(80.93%) | 0.07* |
| Gliclazide | 20230(54.42%) | 893(58.67%) | 171(66.53%) | 0.16* |
| Glimepiride | 4542(12.21%) | 160(10.51%) | 25(9.72%) | 0.03* |
| Metformin | 24235(65.19%) | 993(65.24%) | 168(65.36%) | 0.00* |
| Sulphonylurea | 3486(9.37%) | 121(7.95%) | 16(6.22%) | 0.07* |
| Insulin | 10836(29.15%) | 516(33.90%) | 95(36.96%) | 0.06* |
| Thiozolidinedone | 1644(4.42%) | 49(3.21%) | 8(3.11%) | 0.01* |
| Meglitinide | 3262(8.77%) | 118(7.75%) | 15(5.83%) | 0.08* |
| Glucagon-like peptide-1 agonist | 527(1.41%) | 21(1.37%) | 1(0.38%) | 0.11* |
| Acarbose | 306(0.82%) | 19(1.24%) | 2(0.77%) | 0.05* |
| Diuretics for hypertension | 641(1.72%) | 14(0.91%) | 1(0.38%) | 0.07* |
| Diuretics for heart failure | 713(1.91%) | 43(2.82%) | 19(7.39%) | 0.21 |
| Lipid-lowering drugs | 7130(19.18%) | 144(9.46%) | 32(12.45%) | 0.10* |
| Mean corpuscular volume, fL | 86.82(7.44);134.3;n=17545 | 87.92(7.76);113.4;n=947 | 87.43(7.35);106.9;n=173 | 0.06* |
| Basophil, x10^9/L | 0.04(0.07);7.22;n=12364 | 0.04(0.04);0.2;n=754 | 0.05(0.05);0.2;n=140 | 0.22 |
| Eosinophil, x10^9/L | 0.22(0.25);16.3;n=14131 | 0.22(0.21);1.2;n=829 | 0.26(0.23);1.2;n=156 | 0.15* |
| Lymphocyte, x10^9/L | 2.11(1.0);45.35;n=14136 | 1.78(0.89);6.2;n=830 | 1.92(0.94);5.0;n=156 | 0.16* |
| Metamyelocyte, x10^9/L | 1.4(4.63);22.49;n=23 | 0.11(0.08);0.17;n=2 | - | - |
| Monocyte, x10^9/L | 0.51(0.22);4.14;n=14136 | 0.55(0.25);2.4;n=830 | 0.56(0.25);2.4;n=156 | 0.03* |
| Neutrophil, x10^9/L | 5.03(2.45);72.2;n=14136 | 5.71(2.97);23.5;n=830 | 5.65(2.4);15.9;n=156 | 0.02* |
| White blood count, x10^9/L | 7.88(3.31);180.4;n=17553 | 8.7(7.43);94.6;n=947 | 11.17(16.04);94.6;n=173 | 0.2* |
| Mean cell haemoglobin, pg | 29.2(2.95);43.8;n=17545 | 29.41(3.07);38.9;n=947 | 29.12(2.9);36.1;n=173 | 0.1* |
| Myelocyte, x10^9/L | 0.79(4.17);36.1;n=74 | 0.27(0.13);0.67;n=24 | 0.41(nan);0.41;n=1 | nan |
| Platelet, x10^9/L | 243.18(71.19);783.0;n=17550 | 232.31(84.0);667.0;n=947 | 247.43(106.48);588.0;n=173 | 0.16* |
| Reticulocyte, x10^9/L | 67.9(29.71);175.0;n=234 | 66.26(24.41);103.4;n=25 | 67.99(22.84);99.8;n=10 | 0.07* |
| Red blood count, x10^12/L | 4.66(0.63);9.18;n=17545 | 4.41(0.72);7.23;n=947 | 4.44(0.78);6.9;n=173 | 0.03* |
| Hematocrit, L/L | 0.4(0.05);0.61;n=14383 | 0.38(0.06);0.59;n=783 | 0.38(0.07);0.59;n=135 | 0.01* |
| K/Potassium, mmol/L | 4.31(0.45);8.1;n=28056 | 4.35(0.53);8.1;n=1278 | 4.3(0.48);5.4;n=214 | 0.1* |
| Urate, mmol/L | 0.39(0.11);0.91;n=4475 | 0.42(0.13);0.76;n=186 | 0.47(0.13);0.76;n=47 | 0.41 |
| Albumin, g/L | 42.38(3.57);55.1;n=22334 | 39.82(4.77);50.1;n=1046 | 39.83(4.59);48.6;n=189 | 0.0* |
| Na/Sodium, mmol/L | 139.44(2.81);170.0;n=28063 | 138.83(3.98);170.0;n=1278 | 138.56(4.5);170.0;n=214 | 0.06* |
| Urea, mmol/L | 6.1(2.66);44.6;n=28061 | 7.26(3.94);43.7;n=1277 | 7.38(4.31);36.1;n=214 | 0.03* |
| Protein, g/L | 74.19(5.22);113.0;n=20937 | 72.83(7.16);104.0;n=991 | 73.78(6.49);103.0;n=179 | 0.14* |
| Creatinine, umol/L | 86.54(48.95);1294.0;n=28135 | 108.28(87.33);1276.0;n=1278 | 118.0(124.11);1276.0;n=214 | 0.09* |
| Alkaline phosphatase, U/L | 75.1(28.1);744.0;n=22402 | 87.45(38.25);446.0;n=1051 | 90.17(34.3);286.0;n=191 | 0.07* |
| Aspartate transaminase, U/L | 26.39(27.52);845.0;n=5641 | 31.37(40.1);292.0;n=285 | 30.7(29.29);171.4;n=61 | 0.02* |
| Alanine transaminase, U/L | 30.24(26.78);738.0;n=17686 | 29.96(37.73);303.0;n=815 | 33.0(34.88);182.0;n=141 | 0.08* |
| Bilirubin, umol/L | 11.24(6.21);330.6;n=22296 | 12.17(10.96);152.0;n=1045 | 13.33(12.65);75.0;n=189 | 0.1* |
| Triglyceride, mmol/L | 1.76(1.5);31.0;n=26676 | 1.62(1.03);12.7;n=1127 | 1.66(1.13);12.7;n=191 | 0.04* |
| Total cholesterol, mmol/L | 4.16(1.26);14.8;n=26692 | 4.08(1.4);8.9;n=1128 | 4.15(1.16);7.03;n=192 | 0.05* |
| Low-density lipoprotein (LDL), mmol/L | 2.42(0.82);11.98;n=24428 | 2.42(0.91);6.29;n=1028 | 2.36(0.78);4.54;n=185 | 0.07* |
| High-density lipoprotein (LDL), mmol/L | 1.19(0.32);5.92;n=24929 | 1.17(0.35);3.47;n=1038 | 1.11(0.33);2.31;n=186 | 0.19* |
| Glucose | 8.74(3.83);59.79;n=24502 | 9.35(4.97);51.8;n=1108 | 9.53(5.45);33.49;n=197 | 0.03* |
| HbA1C, g/dL | 11.17(5.25);19.9;n=17808 | 10.83(4.9);19.1;n=956 | 9.96(5.55);19.1;n=173 | 0.17* |
